# Mortality Baselines and Excess Mortality Events in Belgian and Greek Cities Using Singular Spectrum Analysis

**DOI:** 10.64898/2026.10.02.26363997

**Authors:** Damien Delforge, Periklis Charalampous, Natalia Bustos Sierra, Eva De Clercq, Eleni Katragkou, Garyfallos Konstantinoudis, Polychronis Kostoulas, Angeliki Lambrou, Bruno Masquelier, Dimitrios Paraskevis, Wim Thiery, Margo Tonnelier, Niko Speybroeck

## Abstract

**Background:** Excess mortality estimates depend on an expected baseline, an unobservable quantity typically derived from models with prescribed functional forms and short reference periods that exclude exceptional events. Singular Spectrum Analysis (SSA) offers a nonparametric alternative but requires subjective component selection and is sensitive to outliers. We combined SSA with automated component selection and outlier interpolation to estimate long-term baselines and compile a catalog of excess mortality events (EMEs).

**Methods:** We analyzed weekly all-cause mortality (2015–2024) for six metropolitan areas with contrasting climatic, demographic, and epidemiological contexts: Brussels, Antwerp, and Liège (Belgium), and Athens, Thessaloniki, and Larisa (Greece). SSA was applied to standardized log-mortality rates; components were selected using Monte Carlo SSA, and COVID-19 periods were interpolated iteratively. EMEs were defined as weeks exceeding a quasi-Poisson two-z-score threshold, then screened for coincidence with heat waves, cold waves, COVID-19 periods, and influenza epidemics.

**Results:** Baselines were consistent within countries: annual cycles in Belgium, with an additional significant semiannual cycle in Greece reflecting summer mortality. We cataloged 170 EMEs spanning 456 weeks and 14,210 excess deaths. Pandemic weeks accounted for 77% of excess mortality, and 96 weeks coincided with temperature extremes; 56 EMEs, representing 3.7% of excess mortality, remained unmatched.

**Conclusions:** SSA yields reproducible, low-assumption baselines that capture country-specific seasonality. Because flexible baselines can absorb recurrent seasonal mortality, component selection is a substantive choice that should be reported explicitly, together with its sensitivity.

## Background

Excess mortality is defined as a statistically significant deviation from a constant or time-varying expected mortality baseline^1–3^, which itself is an unobservable quantity. It is widely used to monitor and assess the severity of public-health crises and disaster events, including the COVID-19 pandemic,^4–8^ seasonal influenza,^9^ and environmental stressors such as extreme temperatures and air pollution peaks.^10–12^ All-cause excess mortality, more timely available than cause-specific excess mortality, provides a useful framework for surveillance, detection, and investigation of large-scale, possibly compound, events,^13,14^ as illustrated by the EuroMOMO online services (www.euromomo.eu).

As an aggregator, excess mortality captures both direct and indirect causes of death and cannot provide cause-specific attribution, nor the disaggregation prism offered by cause-specific mortality or mortality models with explicit drivers. In surveillance settings, an excess mortality event (EME) is ascribed to whichever anomaly of interest it coincides with (e.g., a heat wave or an influenza epidemic), even though mortality fluctuations are frequently driven by multiple interacting factors.^15–18^ Explicit driver modeling addresses this directly by estimating exposure-lag-response functions,^19^ which capture lagged and nonlinear effects of environmental stressors while adjusting for confounders.^20–23^ The two families answer different questions, with their trade-offs, complementarities, and outcomes being actively discussed, particularly for heat-related mortality.^24,25^ Both, however, depend on an expected baseline, whether explicitly stated or not, and both are sensitive to data resolution and subjective methodological choices.^26–30^

In surveillance practice, this baseline is commonly estimated with log-linear Poisson models adjusted for overdispersion, the excess being quantified from residuals, either standardized (z-scores) or expressed as a percentage of the baseline (p-scores).^2,3^ These models typically include linear and seasonal terms to represent annual cycles,^1,3^ but their prescribed functional forms constrain seasonal patterns and their ability to track long-term demographic changes. Consequently, analyses rely on short reference periods (e.g., five years) and exclude major crises from the baseline estimation,^1–3^ such as the COVID-19 pandemic years (e.g., as in EuroMOMO models). Semi-parametric alternatives based on splines allow greater flexibility over longer time horizons,^31,32^ but can perform poorly for extrapolation and require careful tuning to avoid overfitting.^26^

Nonlinear time-series decomposition methods commonly used in geosciences have been proposed as alternative baseline approaches.^33–36^ Among these, Singular Spectrum Analysis (SSA)—often referred to as Principal Component Analysis for time series—is a non-parametric, data-driven decomposition method that can identify linear, nonlinear, or quasi-periodic patterns in time series data.^34,35,37–39^ SSA therefore offers a low-assumption framework for estimating time-varying mortality baselines over long periods, but raises two practical difficulties: the selection of decomposed components is subjective, and the decomposition is sensitive to extreme outliers, such as COVID-19 pandemic mortality. Both can be addressed using established techniques: Monte Carlo SSA (MCSSA) for component significance testing^40^ and iterative interpolation to obtain robust baseline estimates during extreme events.^41^

The aim of this study is twofold. First, we introduce SSA as an exploratory, low-assumption complement to prescribed-form baselines and implement the practical solutions mentioned above. We focus on two contrasting study areas, three Belgian and three Greek metropolitan areas, whose differing climatic, demographic, and epidemiological contexts make them well-suited to illustrate what an empirical decomposition method like SSA can reveal about temporal patterns in mortality. Second, we extract and catalog EMEs derived from these baselines and further characterize them by their coincidence with temperature extremes and influenza activity. The discussion addresses both the methodological implications of SSA-based baselines and the empirical findings on mortality patterns, EMEs, and their coincidence with temperature extremes and influenza activity.

## Methods

### Data

Weekly all-cause mortality time series, from 2015 to 2024, were obtained for six metropolitan areas from Eurostat.^42^ Data were collected from nine NUTS3 regions, following the European Union’s nomenclature of territorial units for statistics (NUTS, version 2024), corresponding to six metropolitan areas: Brussels, Antwerp, and Liège in Belgium, and Athens, Thessaloniki, and Larisa in Greece (Table 1). The metropolitan areas were selected to represent major urban populations across different European climate contexts (i.e., Belgium and Greece), while also providing variation in population size and mortality patterns within each country. The Athens series aggregates four NUTS3 regions (EL301-EL304), hereafter referred to as EL30X regions. Raw count statistics are shared in the eAppendix (eTable 1). All-cause mortality time series are used in the manuscript, while 80-and-older (80+) mortality data are used as complementary cases in the eAppendix.

**Table 1.** Reference Coordinates and 2015–2024 Mean January 1 Population for Selected NUTS3 Metropolitan Regions.

| **NUTS3 code** | **Metropolitan Areas** | **Latitude<sup>a</sup>**<br>**°N** | **Longitude<sup>a</sup>**<br>**°E** | **Population<sup>b</sup>**<br>***all ages*** |
| --- | --- | --- | --- | --- |
| BE100 | Brussels | 50.84 | 4.39 | 1,224,806 |
| BE211 | Antwerp | 51.28 | 4.50 | 1,061,160 |
| BE332 | Liège | 50.62 | 5.61 | 628,103 |
| EL30X<sup>c</sup> | Athens | 37.99 | 23.75 | 2,608,490 |
| EL522 | Thessaloniki | 40.69 | 23.12 | 1,099,614 |
| EL612 | Larisa | 39.71 | 22.40 | 272,801 |

<sup>a</sup> Latitude and longitude data correspond to centroids extracted from NUTS3 2024 geometries.
<sup>b</sup> Eurostat population data on January 1 by NUTS 3 region, accessed 2026-08-19.<sup>43</sup>
<sup>c</sup> EL30X corresponds to four NUTS3 regions (EL301-EL304).

January 1 population data at the NUTS3 level were obtained from Eurostat,^43^ linearly interpolated, and used to convert counts into rates (Fig. 1). Table 1 provides the mean all-age populations over the study period. Brussels and Athens are fully urbanized areas (eAppendix, eFigure 1); the other regions are predominantly urban with a larger rural share in Larisa (30%). Belgian populations grew over the decade (+2.5% to +7.4%), whereas Greek populations declined (−3.1% to −5.5%), even as their 80+ segment grew, particularly in Thessaloniki and Larisa (+24% and +21%).

**Figure 1.**
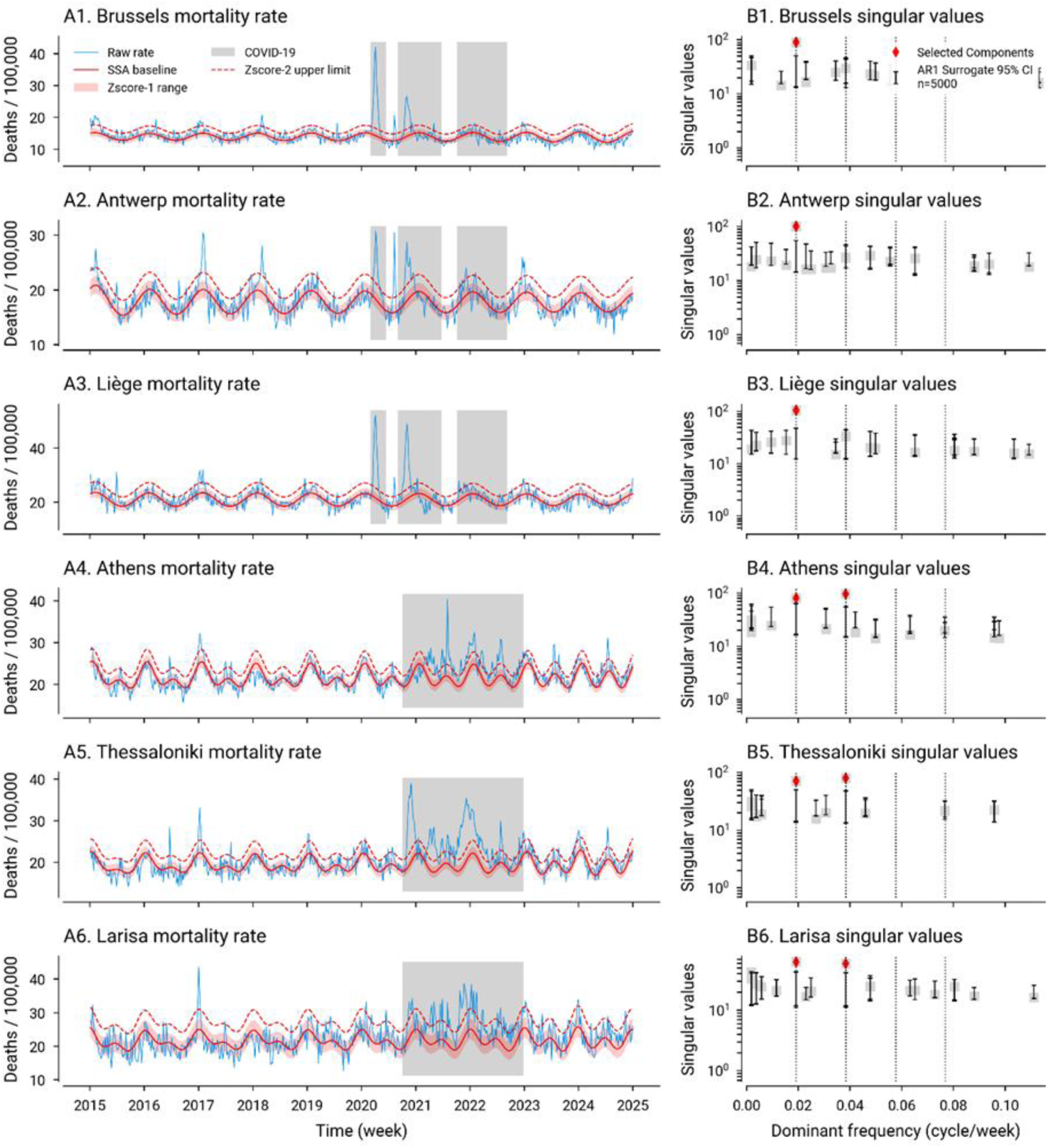
All-Age Mortality Rate Automated SSA Baseline Extraction and Z-Score Envelopes. (A1 to A6) Region-specific mortality time series showing observed data (blue), the SSA-interpolated COVID-19 period (grey), and the automatically SSA-extracted baseline (red) together with the one-z-score envelope and the dashed two-z-score upper threshold. (B1 to B6) Singular values (y-axis) for each region’s SSA decomposition, ordered by the dominant frequencies of the corresponding elementary series (x-axis). Vertical dotted lines indicate annual, six-, four-, and three-month cycles. Significant components exceeding the surrogate-based confidence intervals (red diamonds) correspond primarily to annual cycles in all regions and to semiannual cycles in the Greek regions. SSA was performed using a 261-week window, 5,000 AR(1) surrogates for 95% confidence interval significance testing, and a cutoff frequency of four cycles per year (rightmost vertical line), which had no effect on this baseline extraction (no significant high-frequencies).

Lastly, to assess whether EMEs coincide with extreme temperature events (ETEs), we extracted hourly two-meter air temperature data from the Copernicus ERA5-Land reanalysis,^44^ using the geographic coordinates listed in Table 1. Hourly values in local time were aggregated to obtain daily maximum, mean, and minimum temperatures. The eTables 2-4 summarize the statistical properties of each temperature series for the 1991-2020 climatological reference period used to define heat and cold extremes.

### Mortality Baseline Modeling

Mortality baselines were estimated by applying SSA to z-standardized log-mortality-rate time series, following the univariate approach of Broomhead & King (1986),^45^ as implemented in the SSALib (v.0.1.3) Python package.^46^ SSA is a nonparametric, data-driven method for decomposing a univariate time series into interpretable components, i.e., linear or nonlinear trends, oscillatory patterns (possibly modulated), and noise (residual mortality). We favored this approach over other adaptive methods, such as Empirical Mode Decomposition, because it derives the trend and seasonal cycle from the covariance structure of the series as a whole, rather than relying on its local peaks and troughs.^39^ In our case, this means that individual observations have less influence on the extracted baseline, thereby reducing the risk of inadvertently capturing part of the excess it is intended to separate. Its own requirements—a window length, a choice of components, and the influence that extreme values retain on the decomposition overall—are addressed in the next sections.

Conceptually, SSA extends Principal Component Analysis (PCA) to time series through three steps: (i) construction of a trajectory matrix via time-delayed embedding, Eq. 1, (ii) applying Singular Value Decomposition (SVD, Eq. 2), and (iii) reconstructing the structured signal components.

The first step relies on the window parameter *L*, defining the number of rows in the trajectory matrix **X**. For a time series *X_t_* = {*x*_1_, *x*_2_, …, *x_N_*} of length *N*, the trajectory matrix contains *N* − *L* + 1 lagged segments of the time series, with all anti-diagonal elements being equal.

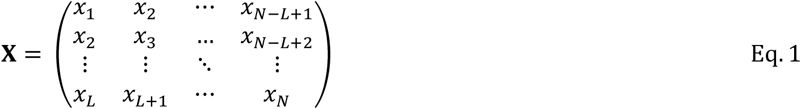

In this study, we set the *L* value to its maximum *N*/2 (261 weeks, or 5 years), a common choice that improves the separability of oscillatory components.^39^ The eAppendix, Section II, further documents the method’s sensitivity to methodological choices.

In a second step, the trajectory matrix was decomposed using SVD (Eq. 2), yielding a sum of rank-one elementary matrices defined by singular values and their associated left and right singular vectors:

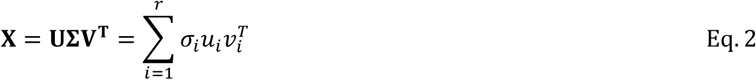

where **U** and **V** hold the left and right singular vectors, **Σ** the singular values in decreasing order, and *r* is the rank of **X**. Each elementary matrix 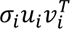 represents a distinct mode of variability, its singular value *σ_i_* reflecting the relative magnitude of the corresponding pattern.

The third step reconstructs a matrix **X***_I_* from a subset of indices *I* = {*i*_1_, *i*_2_, …, *i_k_*} in Eq. 2, which is converted into a time series component 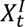 by averaging its anti-diagonal elements (Eq. 1). A singleton *I* yields an elementary series, whose dominant frequency is commonly used to assist component selection.^39^

### Automated Baseline Extraction

To avoid the subjectivity inherent to manual component selection (e.g., based on inspection of singular values, eigenvectors, or dominant frequencies), we automated this step using statistical significance testing through Monte Carlo SSA (MCSSA)^40^, applying a cutoff frequency of 4 yr⁻¹ (i.e., 3-month period) to ensure the extraction of smooth baselines (see eAppendix, eFigure 2). MCSSA uses surrogate data testing to assess the statistical significance of SSA components.^47^ Eigen or singular values are compared with those of synthetic surrogate series, which are typically generated using a first-order autoregressive AR(1) process.^40^ This approach preserves the autocorrelation structure of the observed data (eTable 1). Although AR(1) processes can produce apparent trends or oscillations, these patterns stem from stochastic variability and are transient, making them a suitable null model for significance testing.

Each surrogate series is embedded into a trajectory matrix (Eq. 1) and projected onto the singular vectors obtained from the original SSA decomposition (Eq. 2). The norms of these projections are computed across the surrogate ensemble to construct a 95% confidence interval (95% CI). Singular values from the original SSA decomposition that exceed the surrogate-based CI are considered statistically significant, allowing automated identification of dominant components. MCSSA was implemented using SSALib v.0.1.3.^46^ We also tested the anti-conservative AR(0) white-noise null model in our sensitivity analysis (eAppendix, Section II). Higher-order models had negligible effect on component selection in our cases.

### Baseline Estimation with COVID-19 Outliers

SSA can be sensitive to outliers,^39^ e.g., to the extreme mortality values observed during the COVID-19 pandemic (eFigure 2). To obtain robust baseline estimates in the presence of such outliers, we applied an iterative interpolation approach originally developed for handling missing values in time series.^41^ During the COVID-19 periods, observed values were initially replaced with the mean of the non-COVID-19 period. MCSSA was then applied iteratively, and the COVID-19 period values were updated at each iteration using the statistically significant MCSSA reconstructed baseline components. Iterations continued until convergence, defined as a Root Mean Square Error (RMSE) between two successive interpolations below 10^-3^. The converged reconstruction was retained as the final mortality baseline for subsequent analyses.

Distinct COVID-19 periods were defined for Belgium and Greece. In Belgium, official COVID-19 periods were defined by the national public health institute Sciensano.^48^ These included seven epidemic waves, between March 1, 2020, and September 2022, and two inter-wave periods: from June 22 to August 30, 2020, and from June 28 to October 3, 2021 (Fig. 1). In Greece, COVID-19 mortality followed a more gradual and sustained pattern, without sharply defined waves (Fig. 1). Although the first wave and lockdown began in March 2020, substantial mortality increases were observed primarily from November 2020 onwards.^6,7^ Accordingly, the COVID-19 period for Greece was defined based on visual interpretation as October 1, 2020, to December 31, 2022 (Fig. 1).

### Excess Mortality Events (EMEs)

We back-transformed the MCSSA baseline from the standardized log-rate scale and multiplied it by the weekly population denominator to obtain expected counts µ. Excess mortality was identified as weeks whose observed deaths *Y* reached µ + 2√*φμ*, the two-standard deviation (z-score) limit of a quasi-Poisson variance model, *V*(*Y*) = *φμ*, so the limit widens with the baseline.^2,3,49^ The overdispersion parameter *φ* was estimated per region over non-COVID-19 weeks by iteratively reweighted Pearson residuals to down-weight past excess weeks. As a post-processing step, we applied a one-week-gap tolerance such that two or more events separated by one week with residual mortality below a *z*-score of 2 are considered a single event in our EMEs catalog.

### Extreme Temperature Events (ETEs) Coincidence Analysis

In addition to COVID-19 and seasonal influenza, we examined ETEs, defined as heat and cold waves, that coincided with identified EMEs. We derived ETEs from temperature data and linked them to EMEs by temporal overlap or adjacency. Because ETE definitions vary across studies and regions, depending on variables, minimum durations, intensity thresholds, and reference periods, reported coincidences are sensitive to these parameters.^50–52^

To address this sensitivity, we evaluated a range of candidate ETE definitions per region, defining ETE as periods of minimum duration in which a temperature variable is below (cold) or above (heat) a percentile threshold. Cold-wave thresholds were tested between the 0.5^th^ and 5^th^ percentiles of daily minimum temperature (eTable 3), and heatwave thresholds, between the 95^th^ and 99.5^th^ percentiles of daily maximum temperature (eTable 4). Duration was set to vary between 2 and 6 days.

For each candidate definition, we assessed the coincidence between ETEs and EMEs using the *F*_1_-score, a metric commonly used in binary classification and record linkage,^53,54^ restricted to summer (June–August) for heat waves and winter (December–February) for cold waves. On this basis, we adopted heat waves of at least two days at or above the 99^th^ percentile and cold waves of at least four days at or below the 2.5^th^ percentile (eAppendix, Section II).

## Results

### Mortality Modeling

Figure 1 summarizes the automated MCSSA baseline extraction and excess mortality modeling for each all-age mortality-rate series (Fig. 1A1-6), together with the corresponding singular values ordered by the dominant frequency of their elementary series (Fig. 1B1-6). Analyses used a 261-week window and 5,000 AR(1) surrogates for 95% CI significance testing.

Baseline patterns are consistent within countries. Belgian regions exhibit relatively simple baselines, dominated by annual cycles (Fig. 1B1-3) with mortality peaking during winter months (Fig. 1A1-3), like parametric baselines used in the Be-MOMO or EuroMOMO systems. In contrast, Greek regions show an additional significant semiannual cycle (Fig. 1B4-6), corresponding to increased summer mortality, a pattern that has become more pronounced in recent years (Fig. 1A4-6). A semi-annual cycle was also present in the Belgian series (Fig. 1B1-3), but did not reach statistical significance under MCSSA at the weekly resolution and time span considered.

### Extreme Temperature Events (ETEs) Coincidence Analysis

To identify health-relevant definitions of cold and heat waves, we evaluated the coincidence between ETEs and *z*-score-2 EMEs using the *F*_1_-score framework (Fig. 2). The analysis reveals that a two-day duration combined with the 99^th^ percentile for heat waves resulted in the maximum *F*_1_ (Fig. 2A). While maxima may differ per region, we adopted a common definition considering that the by-region optimums were not significantly different from the all-regions optimum (eAppendix, Section II). Regarding cold waves, many definitions lie on a plateau of near-equivalent choices, and we opted for a middle-ground definition: a daily minimum temperature at or below the 2.5^th^ percentile for at least four days. We refer to the eAppendix for additional details.

**Figure 2.**
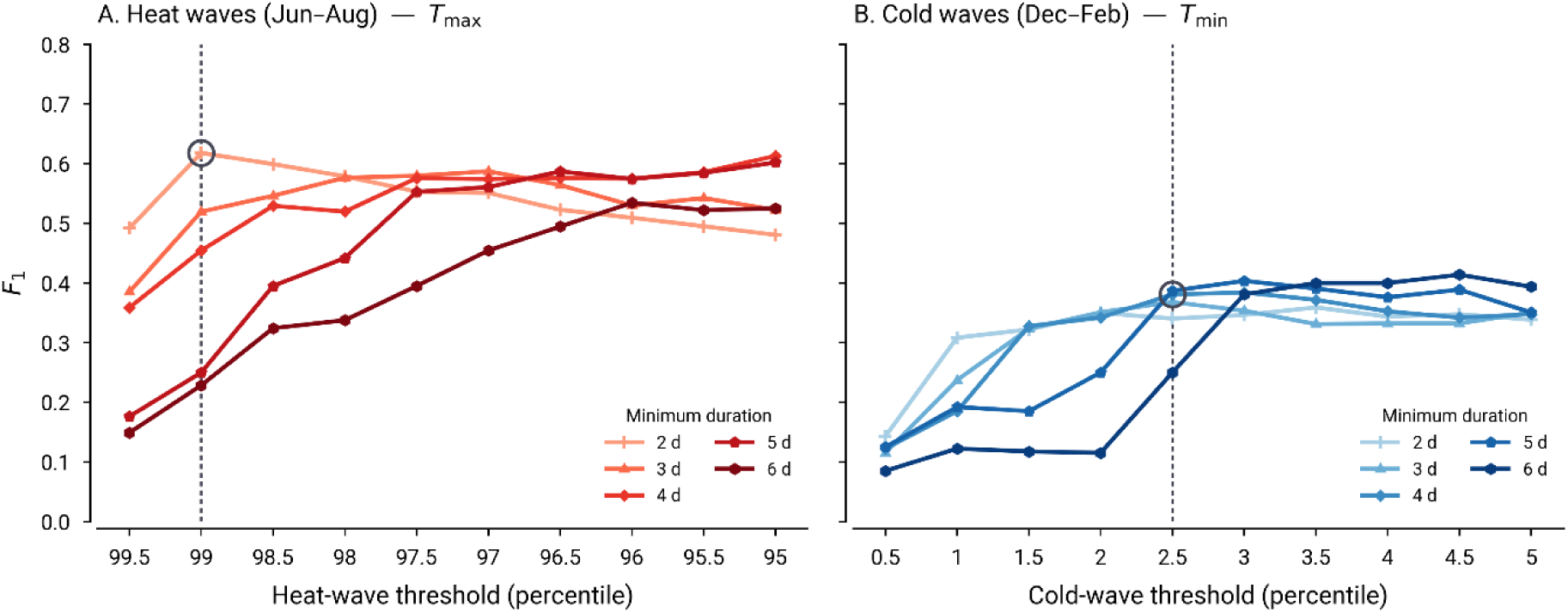
Coincidence Analysis of Z-Score-2 Excess Mortality Events with Extreme Temperature Events Using Various Temperature Percentile Thresholds and Minimum Durations. *F*_1_-scores for heat waves (red, A) and cold waves (blue, B) are shown as a function of temperature percentile thresholds for minimum duration ranging from two to six days. Heat waves are based on daily maximum temperature (*T_max_*) and constrained to summer (Jun–Aug); cold waves on daily minimum temperature (*T_min_*), and constrained to winter (Dec–Feb). The adopted definitions for the subsequent analyses are indicated by the vertical line (adopted percentile) and the circle (adopted duration): heat waves of at least two days at or above the 99^th^ percentile and cold waves of at least four days at or below the 2.5^th^ percentile.

### Excess Mortality Events and Coincident Events

Table 2 summarizes the 170 EMEs identified using a *z*-score threshold of two and a one-week tolerance, spanning 456 excess weeks and 14,210 excess deaths across the six regions. Excess mortality is partitioned by excess weeks, each falling into one of six mutually exclusive categories, crossing the pandemic period (Fig. 1) with identified ETEs. 77% of total excess mortality occurs during pandemic weeks. 96 weeks relate to ETEs, including 30 during the pandemic period. No cold wave coincides with COVID-19 in Belgium.

**Table 2.**
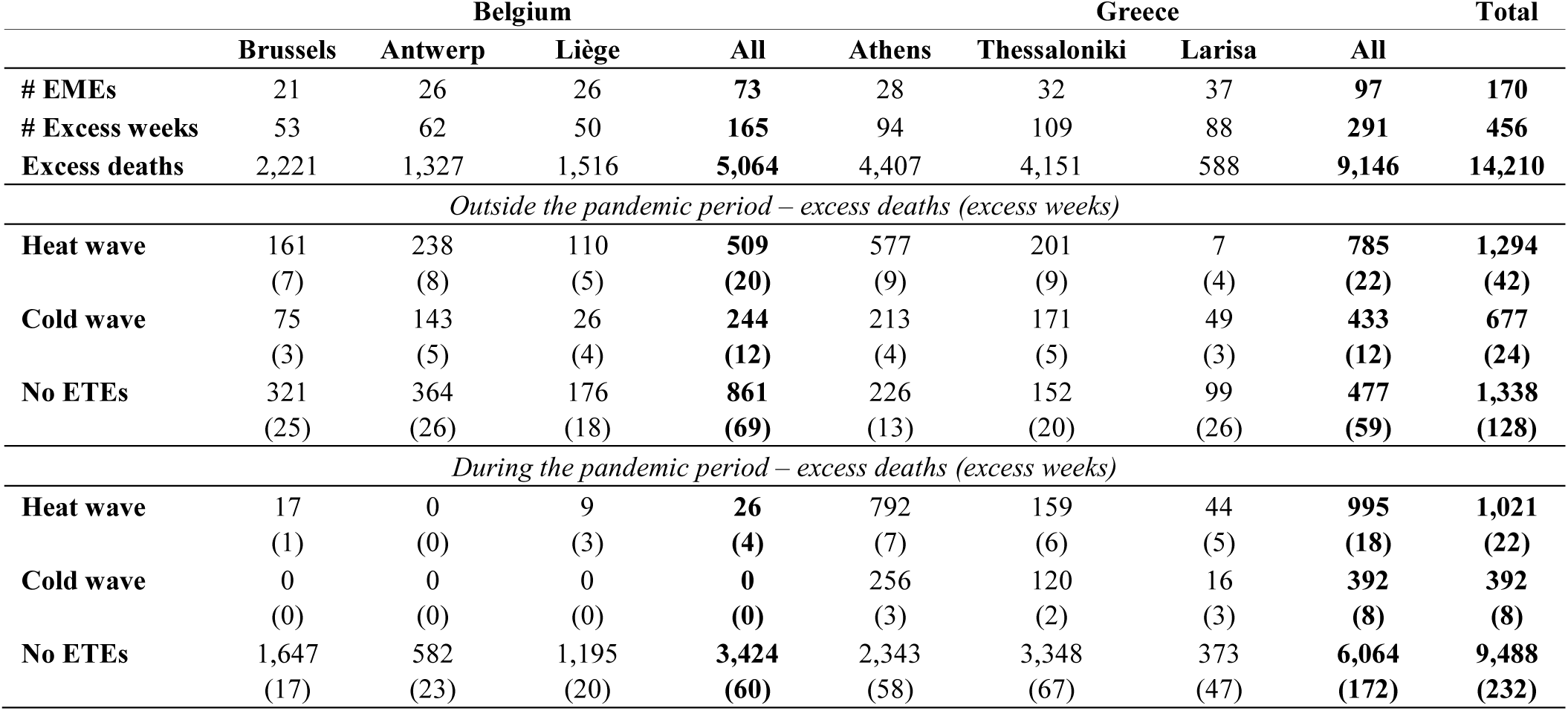
Excess Mortality by Pandemic Period and Coincident Extreme Temperature Events, 2015–2024, Attributed by Weeks.

At the event level, hazards overlap, as shown in Table 3, which lists the top 30 excess mortality events in chronological order. Table 3 also lists coincident influenza epidemics, from country-level reports (eAppendix, Section III). The full catalog of 170 EMEs is provided as a supplement (see Code and Data Availability).

**Table 3.**
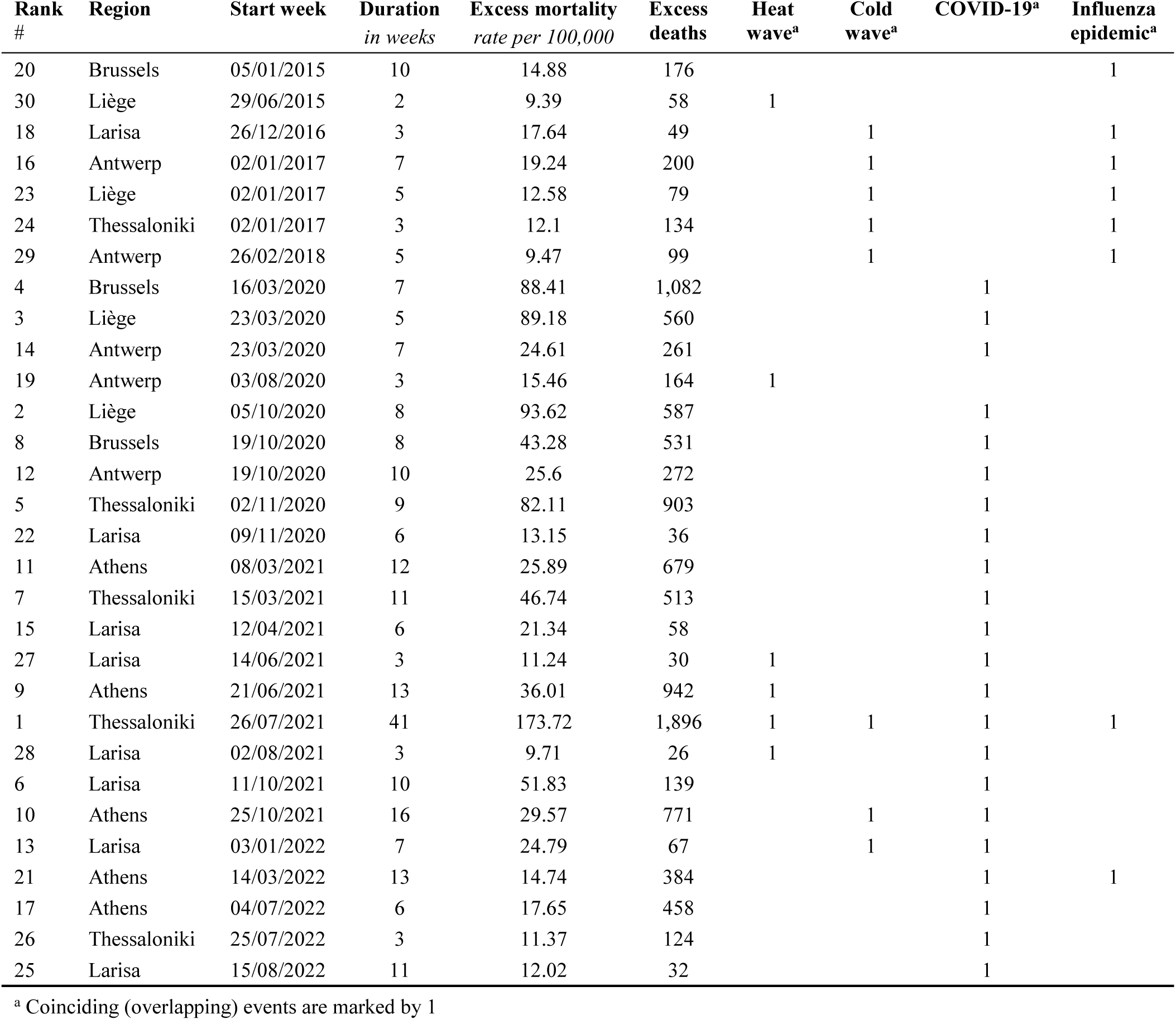
Top 30 Excess Mortality Events (EMEs), 2015–2024, Ranked by Excess Mortality Rate and Presented Chronologically With Their Coinciding Events.

| Rank # | Region | Start week | Duration in weeks | Excess mortality rate per 100,000 | Excess deaths | Heat wave<sup>a</sup> | Cold wave<sup>a</sup> | COVID-19<sup>a</sup> | Influenza epidemic<sup>a</sup> |
| --- | --- | --- | --- | --- | --- | --- | --- | --- | --- |
| 20 | Brussels | 05/01/2015 | 10 | 14.88 | 176 |  |  |  | 1 |
| 30 | Liège | 29/06/2015 | 2 | 9.39 | 58 | 1 |  |  |  |
| 18 | Larisa | 26/12/2016 | 3 | 17.64 | 49 |  | 1 |  | 1 |
| 16 | Antwerp | 02/01/2017 | 7 | 19.24 | 200 |  | 1 |  | 1 |
| 23 | Liège | 02/01/2017 | 5 | 12.58 | 79 |  | 1 |  | 1 |
| 24 | Thessaloniki | 02/01/2017 | 3 | 12.1 | 134 |  | 1 |  | 1 |
| 29 | Antwerp | 26/02/2018 | 5 | 9.47 | 99 |  | 1 |  | 1 |
| 4 | Brussels | 16/03/2020 | 7 | 88.41 | 1,082 |  |  | 1 |  |
| 3 | Liège | 23/03/2020 | 5 | 89.18 | 560 |  |  | 1 |  |
| 14 | Antwerp | 23/03/2020 | 7 | 24.61 | 261 |  |  | 1 |  |
| 19 | Antwerp | 03/08/2020 | 3 | 15.46 | 164 | 1 |  |  |  |
| 2 | Liège | 05/10/2020 | 8 | 93.62 | 587 |  |  | 1 |  |
| 8 | Brussels | 19/10/2020 | 8 | 43.28 | 531 |  |  | 1 |  |
| 12 | Antwerp | 19/10/2020 | 10 | 25.6 | 272 |  |  | 1 |  |
| 5 | Thessaloniki | 02/11/2020 | 9 | 82.11 | 903 |  |  | 1 |  |
| 22 | Larisa | 09/11/2020 | 6 | 13.15 | 36 |  |  | 1 |  |
| 11 | Athens | 08/03/2021 | 12 | 25.89 | 679 |  |  | 1 |  |
| 7 | Thessaloniki | 15/03/2021 | 11 | 46.74 | 513 |  |  | 1 |  |
| 15 | Larisa | 12/04/2021 | 6 | 21.34 | 58 |  |  | 1 |  |
| 27 | Larisa | 14/06/2021 | 3 | 11.24 | 30 | 1 |  | 1 |  |
| 9 | Athens | 21/06/2021 | 13 | 36.01 | 942 | 1 |  | 1 |  |
| 1 | Thessaloniki | 26/07/2021 | 41 | 173.72 | 1,896 | 1 | 1 | 1 | 1 |
| 28 | Larisa | 02/08/2021 | 3 | 9.71 | 26 | 1 |  | 1 |  |
| 6 | Larisa | 11/10/2021 | 10 | 51.83 | 139 |  |  | 1 |  |
| 10 | Athens | 25/10/2021 | 16 | 29.57 | 771 |  | 1 | 1 |  |
| 13 | Larisa | 03/01/2022 | 7 | 24.79 | 67 |  | 1 | 1 |  |
| 21 | Athens | 14/03/2022 | 13 | 14.74 | 384 |  |  | 1 | 1 |
| 17 | Athens | 04/07/2022 | 6 | 17.65 | 458 |  |  | 1 |  |
| 26 | Thessaloniki | 25/07/2022 | 3 | 11.37 | 124 |  |  | 1 |  |
| 25 | Larisa | 15/08/2022 | 11 | 12.02 | 32 |  |  | 1 |  |

<sup>a</sup> Coinciding (overlapping) events are marked by 1

Most EMEs in Table 3 are COVID-19-related (22), ranked among the highest. The Thessaloniki #1 EME is both the deadliest and the longest, spanning 41 weeks, so it coincides with all hazard types. The August 2020 Belgian heat wave, during the COVID-19 inter-wave period (Fig. 1), ranks within the top 30 for Antwerp (#19; #36 and #38 for Brussels and Liège, respectively—see complete catalog for ranks > 30). Greece also experienced heat waves in the summer of 2021 (#27, #9, #1, #28) and cold waves the following winter (#1, #10, #13). In March 2022, Greece experienced a country-wide influenza epidemic (#1, #21, and #32 in Larisa).

Pre-pandemic, 2016-2017 shows a consistent cluster of cold waves and an influenza epidemic, both observed in Belgium (#16, #23, and #41 for Brussels) and Greece (#18, #24, and #35 for Athens). A 2014-2015 influenza epidemic is detected in Brussels (#20), and at lower ranks in other regions (#46 in Antwerp; #66 in Liège). During the 2017–2018 season, a cold wave coinciding with an influenza event is detected in Antwerp (#29), and at lower ranks in Brussels (#43) and Liège (#61). The 30^th^ rank relates to a 2015 heat wave in Liège, also detected in Antwerp (#65) and Brussels (#95).

In general, all events in Table 3 are nationwide or possibly pan-European, such as the COVID-19 pandemic and the 2016-2017 cold waves. Cold waves in Table 3 are either coincident with COVID-19 or influenza events, or both. Across the full catalog, 56 of the 170 EMEs remain unmatched; they account for 3.7% of excess mortality, and 53 of them are single weeks.

## Discussion

### Main Excess Mortality Events

Most of the detected excess mortality is associated with the COVID-19 pandemic (Fig. 1; Tables 2–3). In Belgium, the highest-ranked EMEs clustered around the first two COVID-19 waves of 2020, with all three regions shifting in mid-March (Table 3: #3, #4, #14). Brussels (#4) had 1,082 excess deaths, compared with 1,390 reported during the first wave.^48^ Direct comparison is limited because weekly all-cause, all-age estimates may underestimate wave totals, EME periods do not necessarily align with reported wave periods, and published Belgian estimates are regional rather than city-specific. The record-breaking heat wave in August 2020, during the COVID-19 inter-wave period, was detected in Antwerp (#19) and, with lower death rates, in Brussels (#36) and Liège (#38). Be-MOMO reported 162 excess deaths in Brussels,^55^ compared with 92 in our case, but the peak was particularly acute in Antwerp (#19). Some potential direct or indirect interactions with COVID-19 may have occurred, as reported in different contexts.^17,56,57^

In Greece, excess mortality emerged later than during the March 2020 lockdown, when social-distancing measures largely contained the first epidemic wave.^7,58,59^ Thessaloniki showed a pronounced excess-mortality peak during the autumn 2020 wave (#5), not observed in Athens or Larisa, with previous studies discussing rapid transmission and the city’s role as a major transport hub as possible explanations.^7,60^ Our estimate of 903 deaths is close to the 934 deaths officially registered in Thessaloniki in 2020,^61^ while comparisons for subsequent events are limited by less granular official reporting after November 2020. The greater excess mortality observed in 2021 has been linked, in part, to emerging variants and the burden on healthcare systems, despite the progressive rollout of vaccination.^62,63^

The prolonged COVID-19 EME in Thessaloniki (#1) marginally coincided with the late-January cold wave associated with storm *Elpida* and was also detected in Athens (#10) and Larisa (#13).^64^ It further intersected with the March 2022 countrywide influenza outbreak, ranked #21 for Athens and #32 for Larisa, amid increasing COVID-19 activity; reduced social distancing during carnival festivities has been proposed as a possible explanation.^65^ Lastly, Greece also experienced major summer heat waves during the 2021 pandemic period (#1, #9, #27, #28).^66,67^

Before the pandemic, all Belgian and Greek cities experienced EMEs during the 2016–2017 winter season (e.g., #16, #18, #23, #24 in Table 3; #35, #41 in catalog), coinciding with cold conditions in Greece and across Europe, including Belgium, and a pan-European influenza outbreak.^9,68–70^ The 2014-2015 Belgian high-intensity influenza epidemic—officially lasting 10 weeks and peaking at week 6, 2015—^71^ was detected in Brussels (#20) and, at lower rates, in Antwerp (#46) and Liège (#66), although baseline reference near the start of the study period may be less robust. As a top-ranked ETE at the time,^72^ the 2015 European heatwave EMEs were also captured in late June in Liège (#30) and at lower ranks in Antwerp (#65) and Brussels (#95). Finally, the unusually long 2017–2018 influenza season was associated with EMEs in Antwerp (#29), Brussels (#43), and Liège (#61), coinciding with a cold wave.^73^

Although we cross-checked the largest EMEs in Table 3 against the climate and epidemiological literature, obtaining exact mortality estimates for direct comparison remains challenging. The results nevertheless illustrate the heterogeneous and often overlapping nature of excess mortality, with EMEs coinciding with combinations of pandemic waves, influenza activity, and temperature extremes. This also highlights the value of systematic cataloging: rather than attributing excess deaths to individual hazards, the catalog provides a common framework for identifying and contextualizing mortality anomalies across different settings, which could be pursued forward with explicit mortality driver modeling. While the framework is useful, our catalog estimates should not be interpreted as directly comparable across regions, given differences in population and baseline characteristics.

### Baselines and Excess Mortality: Strengths, Limitations, and Implications

Our results demonstrate that SSA can provide a flexible, reproducible, data-driven approach to constructing long-term mortality baselines that does not rely on a specified functional form. In combination with MCSSA-based component selection,^40^ a low-frequency cutoff, and iterative interpolation of anomalous periods,^41^ the approach produced interpretable baselines across six regions with contrasting mortality patterns. A five-year window (*L* = 261 weeks), consistent with both SSA heuristics^39^ and reference periods used in surveillance,^1–3^ captured the canonical winter-peaking annual cycle in Belgium, while recurrent annual and semi-annual components emerged in the Greek regions. Whether these semi-annual components should be part of the baseline remains purpose-dependent; still, SSA, as an exploratory method, was adequate for highlighting the common pattern across the three Greek metropolitan areas, and the cut-off frequency parameters for SSA components offer a means to control their inclusion in baselines.

The proposed SSA framework addresses practical SSA limitations by enabling automated, reproducible component selection via MCSSA significance testing and dominant-frequency filters, thereby reducing subjective user intervention. This solution fits within a larger body of work on automating SSA,^39^ while the iterative interpolation^41^ provides a pragmatic solution to the influence of extreme mortality events such as COVID-19. Alternative SSA algorithms for incomplete time series exist and may be better suited to other cases.^39,74^ Otherwise, robust alternatives to the classical SVD step, such as weighted or L1-norm formulations,^39,75^ may likewise improve reconstruction in the presence of outliers and could further reduce dependence on ad hoc handling. Our framework is therefore best suited to retrospective, historical reanalysis of mortality as a univariate approach. For real-time surveillance, where forecasting the expected baseline forward is common, established SSA forecasting and multivariate-SSA approaches provide more directly applicable extensions.^39,75^ More generally, as a data-driven approach, SSA may not generalize equally across settings with different data lengths, temporal resolutions, or noise levels; sensitivity testing is therefore recommended before applying the framework elsewhere (see eAppendix).

Our SSA demonstration also has several limitations relevant to interpretation of baselines and EMEs. First, crude rather than age-standardized mortality rates mean that differences in population age structure contribute to differences in baseline levels and EME magnitude, potentially affecting geographical comparisons and rankings (Table 3). Second, NUTS3 aggregates were used as proxies for metropolitan populations, while temperature was represented by gridded values at centroids, which do not perfectly reflect actual exposure conditions. Third, formal uncertainty quantification for expected and excess mortality rates was beyond the scope of this descriptive analysis, which primarily focuses on SSA capabilities, but remains an important direction for future methodological extension.

Fourth, using all-cause mortality aggregates both positive and negative mortality effects, direct and indirect, but does not formally identify their causes. The coincidence analyses with COVID-19, influenza, and temperature extremes should be interpreted as contextual screening rather than attribution. Their purpose is to identify plausible single or overlapping factors that may warrant explicit epidemiological investigation, rather than to establish causal relationships. Such a screening approach is nevertheless valuable because restricting analyses to prespecified drivers can miss concurrent or compound influences on mortality, thereby introducing model specification errors when relevant drivers are omitted. This is further supported by our catalog results (Tables 2 and 3), which highlight frequent compound events. Lastly, EMEs were extracted using different baseline structures for Belgium and Greece; the inclusion of the semi-annual component in the Greek baseline may have resulted in lower estimated summer EME rates by treating part of the recurring summer mortality pattern as expected.

This latter limitation illustrates more broadly why baseline specification is central to the interpretation of excess mortality. Expected mortality is a counterfactual rather than an observable quantity, designed to be fit for purpose.^76^ For instance, whether the Greek summer peaks should be incorporated into expected mortality depends on the intended use of the baseline. From a surveillance perspective, incorporating recurrent seasonal structure may improve specificity for detecting acute departures from prevailing patterns; from a prevention perspective, the same recurring peaks may warrant investigation as potentially preventable heat-related mortality. Baseline components and anomaly-handling decisions should therefore be explicitly reported, rather than remaining implicit within a flexible modeling procedure, together with sensitivity to key modeling choices. These practices would improve the transparency and interpretability of excess mortality estimates and facilitate their comparison across settings.

## Disclaimer

The opinions expressed in this article are those of the authors and do not necessarily reflect the official policy or position of their affiliated institutions, including the National Public Health Organization (NPHO). AL contributed to this work in a personal capacity.

## Data and Computing Code Availability

All data and code sources are available: mortality data (doi:10.2908/DEMO_R_MWEEK3) and population data (doi:10.2908/DEMO_R_PJANGRP3) are available from Eurostat. Temperature data can be accessed through ERA5-land (doi:10.24381/cds.e2161bac). NUTS v2024 tabular data can also be downloaded from Eurostat (URL: https://ec.europa.eu/eurostat/web/nuts). SSA and MCSSA implementation is provided by the Python open-source SSALib package (doi: 10.21105/joss.08600). Output data, including excess mortality and an extreme-temperature event catalog, are available from the Zenodo repository (URL: https://doi.org/10.5281/zenodo.22957843), along with code and a tutorial that enable the reproduction of the baseline results presented in this manuscript.

## Supporting information

eAppendix

