## Supplementary material for "Mortality Baselines and Excess Mortality Events in Belgian and Greek Cities Using Singular Spectrum Analysis": eAppendix

[a] Institute of Health and Society (IRSS), Université catholique de Louvain, Brussels, Belgium.
[b] Department of Epidemiology and Public Health, Sciensano, Brussels, Belgium.
[c] Department of Chemical and Physical Health Risks, Sciensano, Brussels, Belgium.
[d] Department of Meteorology and Climatology, School of Geology, Aristotle University of Thessaloniki, Thessaloniki, Greece.
[e] Grantham Institute for Climate Change and the Environment, Imperial College London, London, United Kingdom.
[f] Laboratory of Epidemiology, Biostatistics and Applied Artificial Intelligence, Faculty of Public and One Health, University of Thessaly, Karditsa, Greece.
[g] National Public Health Organization (NPHO), Athens Greece.
[h] Center for Demographic Research, Université catholique de Louvain, Brussels, Belgium.
[i] Department of Preventive Medicine, Epidemiology and Medical Statistics, Medical School, National and Kapodistrian University of Athens, Athens, Greece.
[j] Department of Water and Climate, Vrije Universiteit Brussel, Brussels, Belgium.

#### Content


I. Data Statistics

Population Statistics

Mortality Statistics

Temperature Statistics

II. Methodological Details

Illustration of MCSSA Applied Without Cut-Off Frequency and Masking COVID-19

Sensitivity of the Mortality Baseline to Modeling and Data Choices

Definition Selection Using Coincidence Analyses Between Excess Mortality and Extreme Temperature Events

III. Additional Results

Excess Mortality per Excess Week

Annual Heatwave and Cold-Wave Trends

Review of Seasonal Influenza Events


**Note.** Additional data and code recipes are available on this online repository: https://doi.org/10.5281/zenodo.22957843

### I. Data Statistics

#### Population Statistics

Population data were downloaded from Eurostat *Population on 1 January by age group, sex and NUTS3 region* dataset (doi:10.2908/DEMO_R_PJANGRP3). The data provide demographic context and insights and are used for estimating mortality rate time series. These population data refer to the selected metropolitan areas described in the main manuscript.

While these areas are primarily metropolitan, the NUTS3 regions may include some rural population. In eFigure 1A, we show the composition of each region using the European Local Administrative Units (LAU) data, version 2024.[1] Each LAU has a population estimate and is classified with a degree of urbanization (DEGURBA) code: (1) cities, (2) towns and suburbs, (3) rural areas. Accordingly, the Brussels (BE100) and Athens (EL301 to EL304) areas can be considered essentially urban. Antwerp, Liège, Thessaloniki, and Larisa have 38%, 30%, 18%, and 16% of their populations living in towns and suburbs, respectively. LAU-based rural population shares are 5% in Liège, 11% in Thessaloniki, and 30% in Larisa.

Total population figures are steadily increasing in Belgium, primarily in Brussels and Antwerp, while the Greek areas show a decreasing trend (eFigure 1B). Population variation relative to 2015 remains contained within ±8%. Regarding the 80+ population, variations are more pronounced, especially in Greece (eFigure 1C). The Greek areas show an increasing 80+ population over 2015–2021, reaching up to 25% above their 2015 level, after which the trend plateaus. In Belgium, Antwerp showed a similar but weaker pattern, increasing over 2015–2020 before leveling off during the COVID-19 and post-COVID-19 periods. For the Brussels and Liège NUTS3 areas, a decrease is observed, accelerating from 2020 onwards. Changes observed in the population pyramids confirm population aging in Greece (eFigure 1D to 1I).

[1] https://ec.europa.eu/eurostat/web/nuts/local-administrative-units, accessed 2026-08-19.

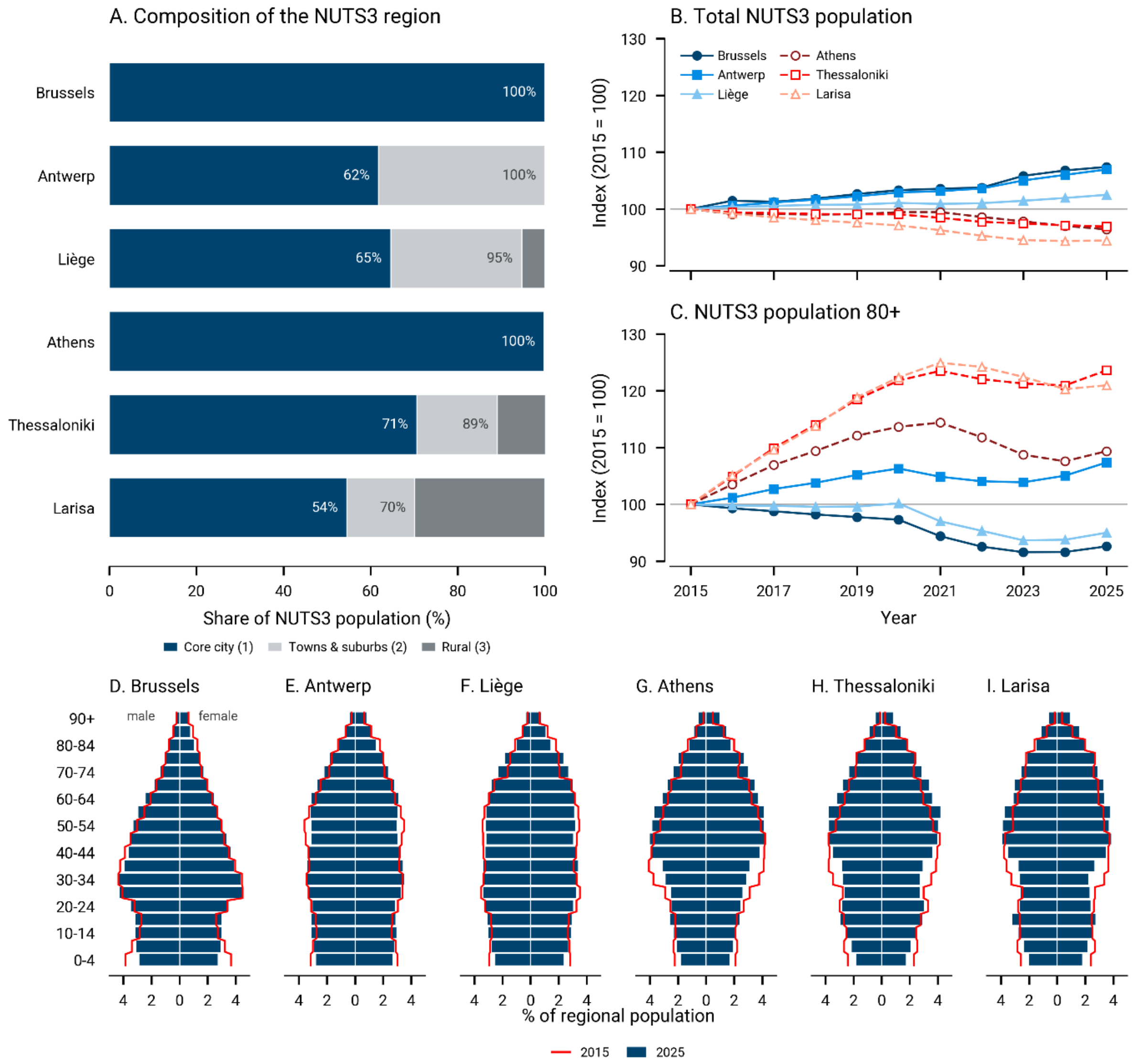


**eFigure 1. Population Data Analysis.** (A) Composition of each NUTS3 region by Eurostat's degree-of-urbanization (DEGURBA) classes based on the Local Administrative Units (LAU) data, version 2024, population statistics, and DEGURBA categories (i.e., urban, suburban, and rural). (B, C) Total and 80+ NUTS3 population temporal dynamics relative to 2015 (= 100), 2015-2025. Belgian regions are drawn with solid lines and filled markers, Greek regions with dashed lines and open markers; marker shape distinguishes the regions within each country. (D to I) Age pyramids of regional populations in 2015 compared with 2025, based on the Eurostat Population on 1 January by age group, sex, *and NUTS3 region* dataset (doi:10.2908/DEMO_R_PJANGRP3).

#### Mortality Statistics

**eTable 1. Statistical Properties of Weekly Mortality Data in Selected Metropolitan Areas (2015-2024, *N* = 522 weeks)**

| | Brussels | Antwerp | Liège | Athens | Thessaloniki | Larisa |
|---|---|---|---|---|---|---|
| *All-age mortality (deaths/week)* | | | | | | |
| **Mean** | 174.9 | 192.0 | 134.9 | 579.1 | 226.2 | 62.6 |
| **Standard deviation** | 35.1 | 30.3 | 26.8 | 77.3 | 40.7 | 12.2 |
| **Minimum** | 120 | 130 | 87 | 409 | 158 | 35 |
| **Percentile 5%** | 140 | 151 | 105 | 479 | 181 | 46 |
| **Percentile 25%** | 155 | 172 | 119 | 527 | 200 | 54 |
| **Percentile 50%** | 169 | 186 | 130 | 565 | 216 | 61 |
| **Percentile 75%** | 186 | 207 | 145 | 614 | 239 | 69 |
| **Percentile 95%** | 224 | 246 | 175 | 726 | 307 | 85 |
| **Maximum** | 515 | 326 | 328 | 1057 | 429 | 121 |
| **Autocorrelation [-]** | 0.75 | 0.69 | 0.73 | 0.74 | 0.78 | 0.53 |
| *80+ mortality (deaths/week)* | | | | | | |
| **Mean** | 98.5 | 117.6 | 72.7 | 347.5 | 133.4 | 39.9 |
| **Standard deviation** | 24.6 | 23.3 | 18.4 | 54.9 | 25.9 | 9.5 |
| **Minimum** | 61 | 67 | 36 | 222 | 80 | 19 |
| **Percentile 5%** | 72 | 87 | 52 | 271 | 98 | 26 |
| **Percentile 25%** | 85 | 101 | 62 | 309 | 116 | 33 |
| **Percentile 50%** | 96 | 114 | 70 | 341 | 129 | 39 |
| **Percentile 75%** | 107 | 130 | 79 | 378 | 146 | 46 |
| **Percentile 95%** | 130 | 162 | 98 | 446 | 183 | 57 |
| **Maximum** | 334 | 232 | 206 | 603 | 262 | 90 |
| **Autocorrelation [-]** | 0.74 | 0.67 | 0.73 | 0.75 | 0.70 | 0.47 |
| *Pearson's correlation coefficient between all-age and 80+ mortality* | | | | | | |
| **Correlation [-]** | 0.95 | 0.95 | 0.94 | 0.96 | 0.93 | 0.90 |

#### Temperature Statistics

**eTable 2. Statistical Properties of ERA5-land Daily Mean Temperature (°C) in Selected Cities (1991-2020, N=10,958)**

| | Brussels | Antwerp | Liège | Athens | Thessaloniki | Larisa |
|---|---|---|---|---|---|---|
| **Mean** | 10.7 | 10.8 | 10.6 | 17.4 | 16.1 | 16.3 |
| **Standard deviation** | 6.4 | 6.3 | 6.6 | 7.4 | 8.3 | 8.2 |
| **Minimum** | -12.1 | -11.6 | -14.0 | -3.2 | -7.5 | -6.8 |
| **Percentile 1%** | -3.4 | -3.2 | -4.0 | 3.3 | 0.2 | 1.4 |
| **Percentile 2.5%** | -1.6 | -1.4 | -2.1 | 4.8 | 1.7 | 2.5 |
| **Percentile 5%** | 0.0 | 0.2 | -0.2 | 6.2 | 3.0 | 3.7 |
| **Percentile 25%** | 6.0 | 6.1 | 5.6 | 11.3 | 9.2 | 9.3 |
| **Percentile 50%** | 10.7 | 10.9 | 10.7 | 17.0 | 16.0 | 15.9 |
| **Percentile 75%** | 15.6 | 15.8 | 15.6 | 24.2 | 23.4 | 23.6 |
| **Percentile 95%** | 20.5 | 20.5 | 20.9 | 28.6 | 28.5 | 29.0 |
| **Percentile 97.5%** | 22.0 | 22.1 | 22.3 | 29.5 | 29.4 | 30.0 |
| **Percentile 99%** | 23.8 | 24.0 | 24.4 | 30.7 | 30.5 | 31.2 |
| **Maximum** | 29.0 | 29.4 | 29.7 | 34.9 | 35.3 | 35.9 |

**eTable 3. Statistical Properties of ERA5-land Daily Minimum Temperature (°C) in Selected Cities (1991-2020, N=10,958)**

| | Brussels | Antwerp | Liège | Athens | Thessaloniki | Larisa |
|---|---|---|---|---|---|---|
| **Mean** | 7.2 | 7.5 | 7.2 | 13.1 | 12.2 | 11.9 |
| **Standard deviation** | 5.8 | 5.8 | 6.1 | 6.5 | 8 | 7.4 |
| **Minimum** | -13.5 | -14.1 | -17.1 | -7.7 | -11.5 | -11.7 |
| **Percentile 1%** | -5.8 | -5.6 | -7.1 | 0.7 | -3.1 | -1.5 |
| **Percentile 2.5%** | -4.2 | -3.9 | -4.7 | 1.9 | -1.5 | -0.4 |
| **Percentile 5%** | -2.7 | -2.4 | -3 | 3.1 | -0.5 | 0.7 |
| **Percentile 25%** | 2.9 | 3.2 | 2.8 | 7.7 | 5.4 | 5.7 |
| **Percentile 50%** | 7.5 | 7.7 | 7.5 | 12.9 | 12.1 | 11.5 |
| **Percentile 75%** | 11.8 | 12.1 | 11.9 | 19.1 | 19.3 | 18.5 |
| **Percentile 95%** | 15.9 | 16.1 | 16.3 | 22.9 | 24.2 | 23.1 |
| **Percentile 97.5%** | 17.2 | 17.3 | 17.5 | 23.6 | 25.2 | 24 |
| **Percentile 99%** | 18.3 | 18.6 | 19 | 24.4 | 26.1 | 25 |
| **Maximum** | 22.5 | 23.2 | 24.4 | 27.4 | 30.4 | 27.9 |
| **Correlation**[a] | 0.965 | 0.966 | 0.967 | 0.976 | 0.988 | 0.983 |

[a] Pearson's correlation coefficient between Daily Mean Temperature (eTable 2) and Daily Minimum Temperature

**eTable 4. Statistical Properties of ERA5-land Daily Maximum Temperature (°C) in Selected Cities (1991-2020, N=10,958)**

| | Brussels | Antwerp | Liège | Athens | Thessaloniki | Larisa |
|---|---|---|---|---|---|---|
| **Mean** | 14.2 | 14.2 | 14 | 21.9 | 20.2 | 20.9 |
| **Standard deviation** | 7 | 7 | 7.3 | 8.3 | 8.6 | 9 |
| **Minimum** | -9.9 | -9.4 | -11.9 | -0.6 | -5.9 | -2 |
| **Percentile 1%** | -0.8 | -0.6 | -1.2 | 5.4 | 2.8 | 3.5 |
| **Percentile 2.5%** | 1.2 | 1.1 | 0.6 | 7.3 | 4.8 | 5.4 |
| **Percentile 5%** | 2.9 | 2.9 | 2.4 | 8.9 | 6.5 | 7 |
| **Percentile 25%** | 8.9 | 8.9 | 8.5 | 15.1 | 13.2 | 13.6 |
| **Percentile 50%** | 14.3 | 14.2 | 14 | 21.5 | 20.2 | 20.7 |
| **Percentile 75%** | 19.5 | 19.4 | 19.5 | 29.2 | 27.7 | 28.8 |
| **Percentile 95%** | 25.7 | 25.7 | 25.8 | 34.6 | 33.2 | 34.6 |
| **Percentile 97.5%** | 27.5 | 27.5 | 27.7 | 35.8 | 34.3 | 36.1 |
| **Percentile 99%** | 29.8 | 30 | 29.9 | 37.3 | 35.6 | 37.7 |
| **Maximum** | 37.5 | 38.3 | 37.9 | 43.9 | 41.2 | 43.9 |
| **Correlation**[a] | 0.981 | 0.981 | 0.982 | 0.988 | 0.991 | 0.987 |

[a] Pearson's correlation coefficient between Daily Mean Temperature (eTable 2) and Daily Maximum Temperature

#### II. Methodological Details

##### Illustration of MCSSA Applied Without Cut-Off Frequency and Masking COVID-19

In eFigure 2, MCSSA is applied to the log-mortality-count time series of Brussels without a cut-off frequency and without masking the COVID-19 period (eTable 1). In this case, the automatically MCSSA-extracted signal exhibits a peculiar seasonal pattern (eFigure 2A), as it includes significant components (red diamonds, eFigure 2C) with corresponding frequencies near 4- and 6-month periods, though not perfectly aligned. These spurious harmonics most likely result from the COVID-19 outliers, such that the extracted signal phases align with observed COVID-19 peaks (eFigure 2A). For this reason, baselines for the COVID-19 periods are interpolated for the results shown in the main manuscript.

In addition, the MCSSA extracted high-frequency variations at ~0.29 weeks$^{-1}$ (approx. 3.5 weeks) in eFigure 2D, corresponding to indices 36 and 37 in eFigure 2C. Such low-variance and high-frequency variations are excluded in the main manuscript by setting a cut-off frequency of 4/52 week$^{-1}$, corresponding to the 3-month period in eFigure 2D.

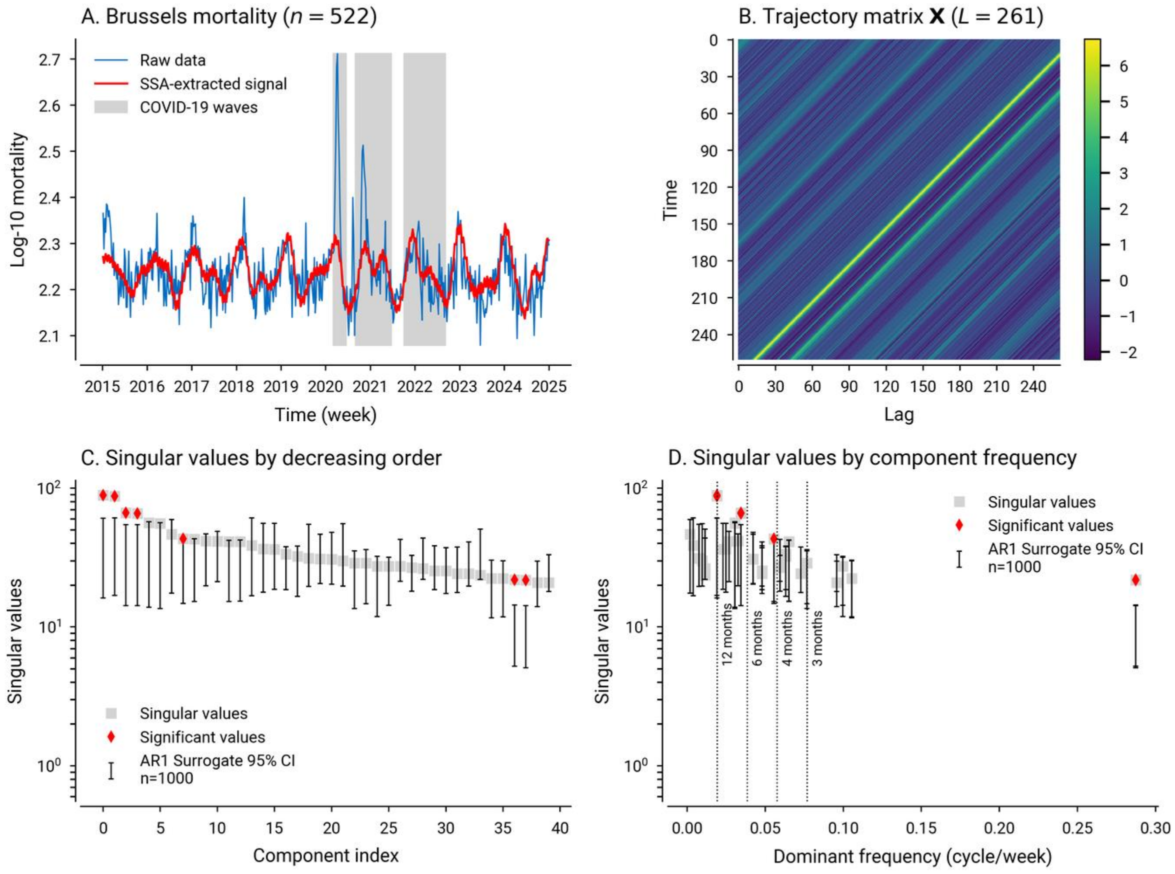


**eFigure 2. Monte Carlo SSA Decomposition of the Brussels Log-Mortality Time Series, Problematic Due to COVID-19 Outliers.** (A) Raw time series (blue) and signal automatically reconstructed using MCSSA (red), with the COVID-19 period in the background (grey); (B) Trajectory matrix associated with the z-standardized log-mortality series using an embedding window of 261 weeks; (C) Singular values by decreasing order (grey squares) and surrogate projections (error bars), with highlighted significant values (red diamonds); (D) Same as (C) ordered by component dominant frequency, with frequencies associated with 3 to 12 month periods (vertical dotted lines). This illustration uses 1,000 AR(1) surrogates.

Replicates of eFigure 2 for the other metropolitan areas are available in the online repository. As in eFigure 2, they decompose weekly death counts rather than the mortality rates used for event extraction and are therefore uncorrected for population change (eFigure 1), which favors low-frequency components (near-zero frequencies in panel d). These are strongest in the Greek series, where COVID-19 also produced smoother and more sustained increases in mortality than the acute waves seen in Belgium.

#### Sensitivity of the Mortality Baseline to Modeling and Data Choices

To support our modeling choices and analyze their sensitivity, we repeated the baseline extraction over a full factorial grid of four binary choices, presented in eTable 5. The remaining parameters were held fixed: the cut-off frequency corresponding to a 3-month period and the number of surrogates (n=5,000), set intentionally high to increase the robustness of the analysis. Regarding the MCSSA, we compare only two surrogate null models: $AR(0)$ and $AR(1)$. Increasing the order above 1 could yield a better model according to the Bayesian Information Criterion (BIC); however, it did not change the selected components compared with the $AR(1)$ model and, consequently, did not affect the subsequent results.

The modeling choice grid yields 16 baselines for each metropolitan area, all of which can be accessed in the online repository. The reference case — marked by a footnote letter in eTable 5 — corresponds to Fig. 1 in the main manuscript.

**eTable 5. Modeling Choices Explored in the Sensitivity Analysis**

| # | Modeling Choice | Option 1 | Option 2 |
|---|---|---|---|
| 1 | Decomposed quantity | Weekly death counts | Weekly mortality rates[a] |
| 2 | Embedding window | $L = 130$ weeks (2.5 years) | $L = 261$ weeks (5 years)[a] |
| 3 | MCSSA surrogate null | White noise ($AR(0)$) | Red noise ($AR(1)$)[a] |
| 4 | Population group | Total (all ages)[a] | 80+ |

[a] Modeling choices used as reference in the main manuscript.

This sensitivity analysis specifically targets the question: "*For a given population group (#4), how different would the reference catalog be if one changes modeling choices #1 to #3 one at a time?*" Instead of comparing segmented events, we set the unit to excess-mortality weeks. This unit avoids confusion that can arise from comparing events of different durations and varying overlaps. As a comparison metric, we opted for the Jaccard index, i.e., the intersection over the union of two sets $A$ and $B$:

$$J = |A \cap B| / |A \cup B| \qquad Eq. S1$$

where one of the two sets is the fixed reference case (eTable 5, #1-3, Option 2). Weeks flagged by neither specification, about 90% of the record, are excluded by construction, so $J$ is not inflated by agreement on the quiet majority of the series. $J$ is therefore an indicator of the degree of overlap between excess-mortality-week sets, and is used here to address the sensitivity reported in eFigure 3.

Specifically, eFigure 3 reports, for each modeling choice #1–3 (y-axis) and population group (panels a vs. b), the mean $J$ across the six metropolitan areas relative to the reference case (bars). Individual areas are also reported, each with its own marker, colored by country. We make three main observations for the total population (eFigure 3A). First, the surrogate null effect dominates in the total population group ($J$ = 0.67) and the 80+ population ($J$=0.78, tied with the decomposed quantity). A white-noise null is anti-conservative for a strongly autocorrelated series and overly permissive in component selection: trend and low-frequency variation are more readily admitted into the baseline—particularly for mortality counts, which are influenced by population dynamics—along with other frequencies that are difficult to justify. eFigure 4 illustrates this for the rate-based baselines and shows components retained well beyond the annual and semiannual cycles identified in Fig. 1.

Second, the embedding window has a moderate effect on average on the total population ($J$ = 0.83), but one carried by two regions: Athens (0.69) and Larisa (0.59) against 0.90 to 0.93 elsewhere. In this case, Athens shows a baseline reduced to the semiannual cycle, and Larisa shows one composed of a trend and an annual cycle. For the 80+ population, Larisa's baseline only includes an annual cycle, with the resulting $J$ lying below the average. Nonetheless, a window of 261 weeks corresponds to $N/2$ and to five annual cycles and remains the SSA default chosen in the main manuscript to optimize cycle detection. The $N/4$ window of 130 weeks can also be considered contentious, as it is shorter than the COVID-19 period.

Third, changing the decomposed quantity from rates to counts has limited effects on total population ($J$ = 0.92), the mildest of the three perturbations (eFigure 3A). Results are more dispersed in the 80+ population panel (eFigure 3B), and the three choices no longer rank as they do for the total population. The decomposed quantity becomes as consequential as the surrogate null ($J$ = 0.78 for both), and it is here that the Greek metropolitan areas are the most sensitive — Larisa 0.54, Thessaloniki 0.70, and Athens 0.78, against 0.79 to 0.97 in the Belgian regions. This is consistent with the greater observed changes in this population group (eFigure 1C): the 80+ denominator moves by up to 25% over the study period, against at most 7% for the total population, so dividing by it matters far more. The surrogate null behaves in the opposite way: Athens and Thessaloniki are unaffected ($J$ = 1.000, i.e. no flagged week changes), while the three Belgian regions move furthest (0.63 to 0.65). Overall, Larisa — also the least-populated area — is the most sensitive region wherever the seasonal structure of the baseline is at stake.

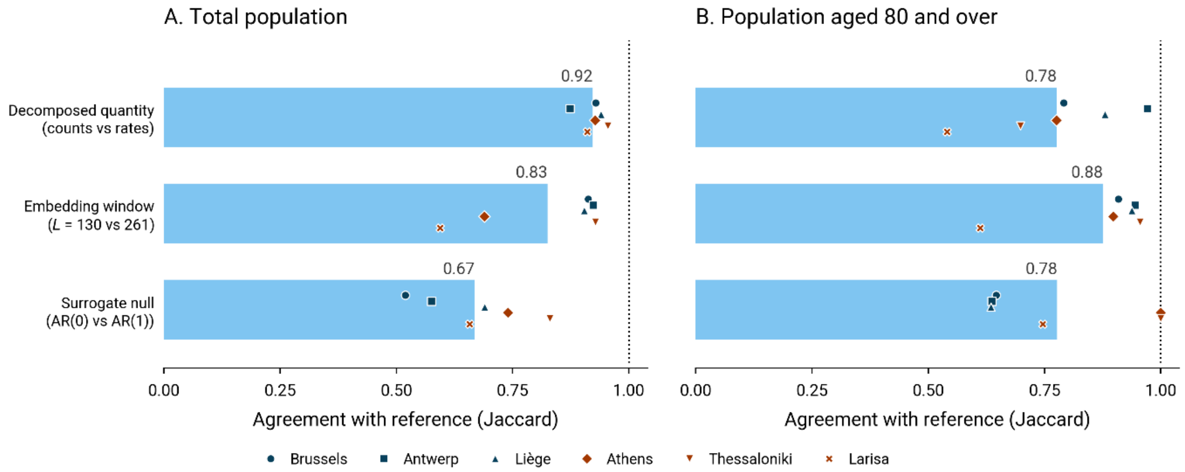


**eFigure 3. Sensitivity of the Detected Excess Mortality Weeks to the Three Modeling Choices Behind the Mortality Baseline.** Each bar reverses one choice from the reference specification (eTable 5: mortality rates, L = 261 weeks, AR(1) surrogate null) while holding the other two at the reference, and gives the Jaccard index of flagged weeks between the two specifications — the share of weeks flagged by either that are flagged by both — averaged over the six metropolitan areas. Markers correspond to individual metropolitan areas, in blue for Belgium and orange for Greece. The dotted line marks the perfect agreement the reference attains with itself, so every bar is a reduction from it. (A) Total population. (B) Population aged 80 and over, scored against its own reference, in the factor order of panel (A).

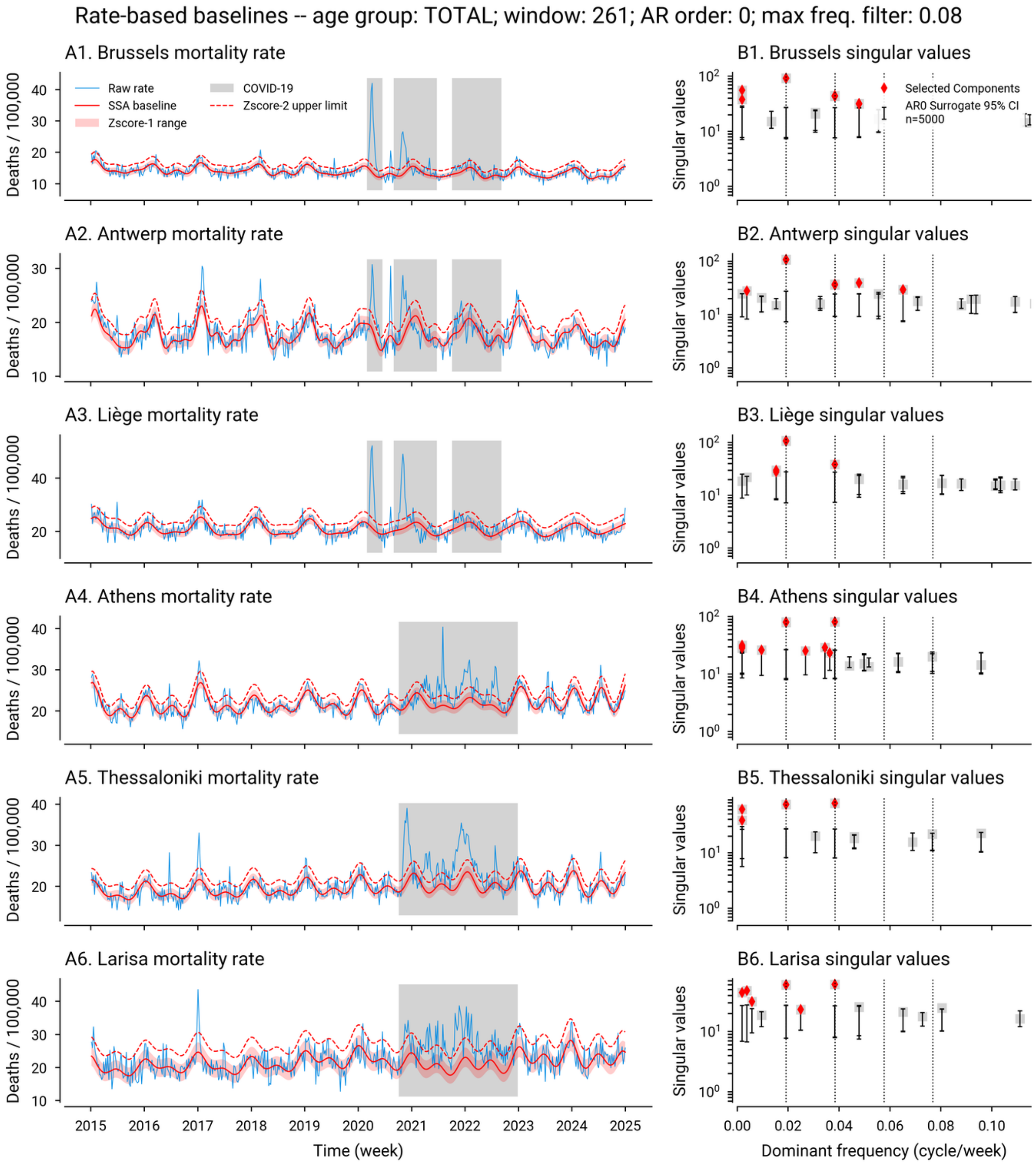


**eFigure 4. All-Age Mortality Rate Automated SSA Baseline Extraction and Z-Score Envelopes Using a White-Noise (AR(0)) Surrogate Null.** This is the non-reference illustration from the sensitivity grid; the reference case, which differs only in its AR(1) surrogate null, is Fig. 1 of the main manuscript. (A1 to A6) Region-specific mortality rate time series (deaths per 100,000) showing observed data (blue), the SSA-interpolated COVID-19 period (grey), and the automatically SSA-extracted baseline (red) together with the one-z-score envelope and the dashed two-z-score upper threshold. (B1 to B6) Singular values (y-axis) for each metropolitan-area SSA decomposition, ordered by the dominant frequencies of the corresponding elementary series (x-axis). Vertical dotted lines indicate annual, six-, four-, and three-month cycles. Red diamonds correspond to significant components exceeding the surrogate-based confidence intervals; only those to the left of the cutoff frequency enter the baseline. SSA was performed using a 261-week window and 5,000 AR(0) surrogates for 95% confidence interval significance testing, with a cutoff frequency of four cycles per year (rightmost vertical line).

**Definition Selection Using Coincidence Analyses Between Excess Mortality and Extreme Temperature Events**

*Method Description*

We define Extreme Temperature Events (ETEs) as runs of consecutive days, of at least a minimum duration, on which a daily temperature statistic lies beyond a percentile threshold of the 1991–2020 daily distribution, computed separately for each metropolitan area. Operational definitions vary in both percentile and duration across regions, populations, and purposes, and no external standard settles the choice for six areas spanning two climates. In a mortality-focused study, however, we would expect a relevant heatwave (HW) or cold-wave (CW) definition to show some coincidence, or overlap, with Excess Mortality Events (EMEs). We therefore selected the definition on that criterion, applying one definition to all six areas and scoring it on their pooled coincidence with excess mortality.

This section focuses on the percentile and duration parameters of the ETE definitions. We also investigated two other aspects and did not include them in the figures and tables below to simplify reporting: the underlying temperature variable (i.e., min, max, mean), and whether percentiles should be defined based on the full-year temperature distribution or on calendar-day windows. We concluded that (i) defining heat waves on daily maximum temperatures (eTable 4), (ii) cold waves on daily minimum temperatures (eTable 3), and (iii) using full-year climatology provided the best coincidence with EMEs. The percentiles reported below are therefore fixed temperature thresholds for each metropolitan area — as opposed to the seasonally varying thresholds often used in the climate literature — and can be read in °C (from eTable 3 and 4).

Below, we report a coincidence analysis over a grid of 10 percentiles per hazard and five minimum durations (two to six days), yielding fifty candidate definitions per hazard. Candidate definitions were scored against the excess mortality catalog described in the main manuscript. We considered event adjacency (no overlap but directly consecutive events) as a coincident match when ETEs preceded EMEs, on the reasoning that extreme temperature precedes mortality. Scoring was also restricted by season: each EME was assigned the meteorological season of the week in which it began, and heat waves were scored only against the 62 summer (Jun-Aug) EMEs and cold waves only against the 44 winter EMEs (Dec-Feb), out of the 170 EMEs detected across all seasons of the 2015-2024 study period. Coincidence was then summarized by precision ($P$), recall ($R$), and their harmonic mean, the $F_1$ score. Writing $E$ for the ETEs of one hazard and $M$ for the EMEs of its season, and $E^*$ and $M^*$ for those members of each set linked to at least one event of the other:

$$P = \frac{TP_E}{TP_E + FP} \qquad Eq. S2.1$$

$$R = \frac{TP_M}{TP_M + FN} \qquad Eq. S2.2$$

$$F_1 = \frac{2}{P^{-1} + R^{-1}} \qquad Eq. S2.3$$

where $TP_E$ is the number of temperature events linked to at least one excess mortality event and $FP$ the number linked to none; $TP_M$ is the number of excess mortality events linked to at least one temperature event and $FN$ the number linked to none. Precision $P$ (or positive predictive value) is thus the proportion of temperature events coinciding with excess mortality, and recall $R$ (or sensitivity) the proportion of seasonal excess mortality events coinciding with a temperature event.

Precision, recall, and $F_1$ are reported as computed on the full sample. We used the bootstrap only to obtain 95% confidence intervals, not to re-estimate the statistics. The resampling unit is the metropolitan-area season-year: all events falling in one area in one season, i.e., summer (heat) or winter (cold). Blocks are drawn with replacement, as many as the sample contains (52 area-summers for heat waves and 44 area-winters for cold waves under the adopted definitions); both catalogs are restricted to the drawn blocks, and the metrics are recomputed; the intervals are the 2.5$^{th}$ and 97.5$^{th}$ percentiles of 4,000 such resamples.

*Coincidence Analysis and Selected Definitions*

eFigure 5 reports $F_1$, $P$, and $R$ (rows) against the percentile threshold, with one line per minimum duration and the two hazards side by side (columns); the adopted definition is ringed. We see a flat $F_1$ curve conceals two large opposing movements: a decreasing $P$ and increasing $R$. Across the heatwave grid (panels A, C, E), $P$ falls from 1.00 at the strictest combination to 0.33 at the loosest while $R$ rises from 0.08 to 0.87, leaving $F_1$ almost unchanged over much of the range. The confidence intervals are wide and overlap extensively, so most candidate definitions are statistically indistinguishable from the best-performing one.

The adopted definitions are *heat waves of at least two consecutive days with a daily maximum temperature at or above the 99$^{th}$ percentile*, and *cold waves of at least four consecutive days with a daily minimum temperature at or below the 2.5$^{th}$ percentile*. eTable 6 reports what they yield. Over the six areas, 50 of 78 heat waves coincide with an excess mortality event and 37 of 62 summer excess mortality events with a heat wave, giving $F_1$ = 0.62; for cold waves the corresponding counts are 15 of 28 and 13 of 44, giving $F_1$ = 0.38. The adopted heatwave definition is the highest-scoring of the fifty; the adopted cold-wave definition scores 0.03 below the best-scoring cell and ranks eleventh of fifty, a difference far smaller than the width of either confidence interval. The definitions should therefore be read as lying on a plateau of near-equivalent choices rather than as a sharp optimum, and were selected within that plateau to balance precision and recall and to maintain consistency with conventional thresholds.

Still, definition selection criteria remain context-dependent: an F-measure weighted toward recall could be considered in public health emergency or disaster risk management contexts, on the reasoning that missing a relevant event is more costly than a false alarm. However, such a criterion would not be informative on this grid, because recall cannot decrease as the definition is loosened: loosening an intensity threshold or shortening a minimum duration can only add exceedance days, so the events detected under a looser definition always contain those detected under a stricter one, making recall monotonically non-decreasing across the grid by construction. Any criterion that favors recall strongly enough therefore selects the loosest definition on the grid, regardless of the data, and cannot discriminate among candidates.

Regarding hazard-specific patterns, coincidence is weaker for cold waves than for heat waves throughout the grid. Excess winter mortality has competing drivers, seasonal influenza in particular, that do not necessarily coincide with cold spells, so a larger share of winter excess mortality has no associated temperature event. This shows in recall rather than in precision: cold waves that do occur are about as likely as heat waves to coincide with excess mortality ($P$ 0.54 against 0.64), but far more excess mortality occurs without them ($R$ 0.30 against 0.60). The heatwave results also differ between the two countries, with the same number of heat waves — 39 in each — and higher scores in the Greek areas than in Belgium ($F_1$ = 0.67 vs 0.54; eTable 6), with the difference lying in precision (0.79 vs 0.49) rather than recall.

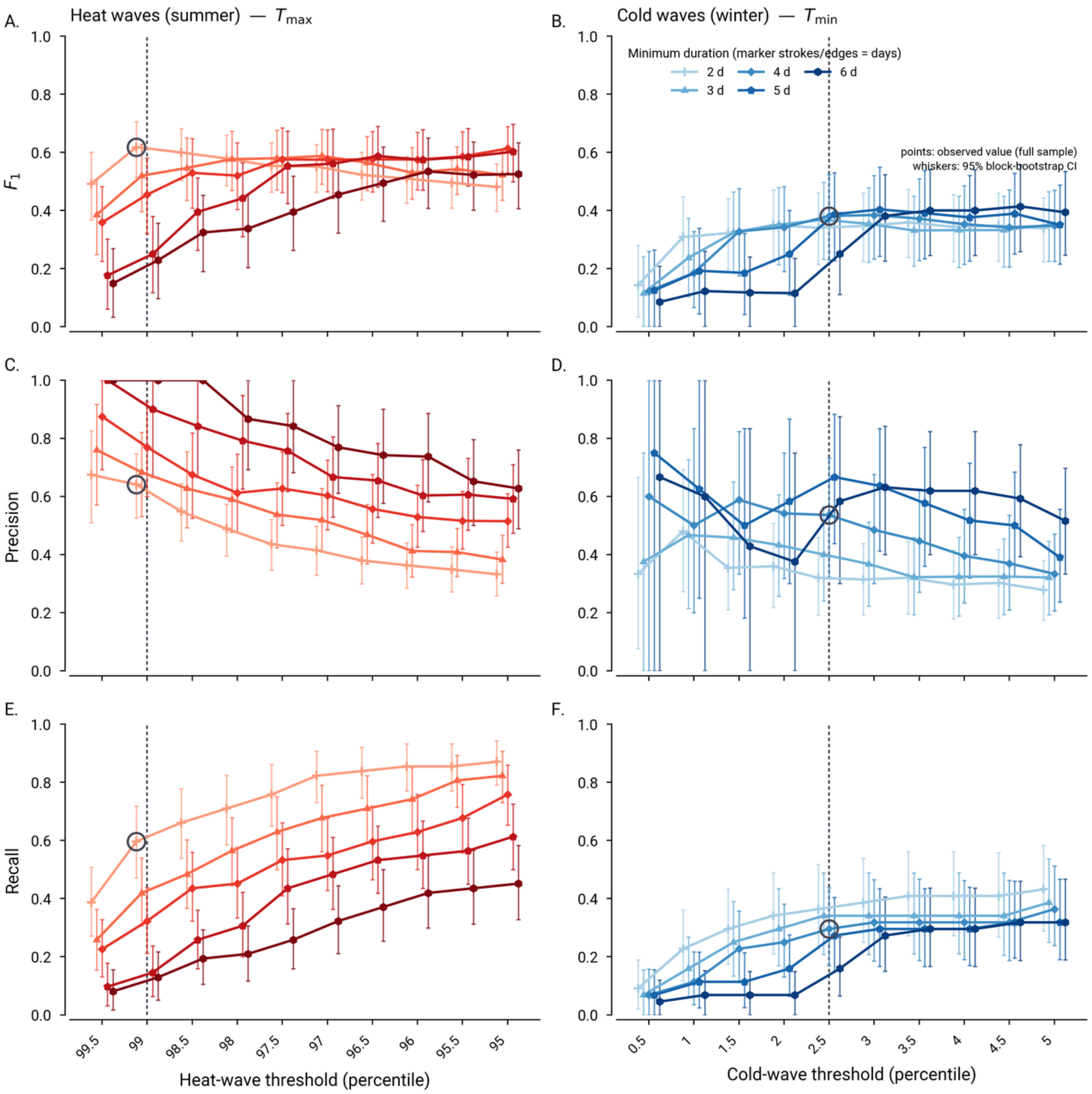


**eFigure 5. Coincidence Analysis Between Extreme Temperature Events and Excess Mortality Events Across Candidate Definitions.** Each panel plots one metric against the definition's percentile threshold, with one line per minimum duration (two to six days; the number of strokes or edges of the marker indicates the duration in days). (A, C, E) Heat waves, scored on daily maximum temperature against summer excess mortality events. (B, D, F) Cold waves scored on daily minimum temperature against winter excess mortality events. (A, B) $F_1$ score (Eq. S2.3); (C, D) precision (Eq. S2.1); (E, F) recall (Eq. S2.2). Thresholds are percentiles of the 1991–2020 daily distribution computed for each metropolitan area, and become less extreme from left to right in both columns. Points are the values observed in the full sample; whiskers are 95% percentile bootstrap intervals from 4,000 resamples of metropolitan-area season-years. The dashed vertical line marks the adopted threshold and the ringed marker the adopted definition: heat waves of at least two days at or above the 99th percentile of daily maximum temperature, cold waves of at least four days at or below the 2.5th percentile of daily minimum temperature.

Because the adopted definition is selected on the pooled catalog, a pooled optimum could conceal a difference between the two countries. eFigure 6 rescores the same grid separately for the Belgian and the Greek metropolitan areas. For heat waves, the two surfaces differ in shape: coincidence in Greece is sharply peaked and its maximum is the adopted definition itself, whereas in Belgium it is flat across a broad range of thresholds and its nominal maximum falls at the 95th percentile over at least four days. That Belgian maximum lies on the lowest threshold the grid sweeps, so it marks a plateau rather than a distinct optimum, and it is not statistically distinguishable from the adopted definition, with each point estimate falling within the other's confidence interval. Cold-wave coincidence is flat above roughly the third percentile in both countries. We therefore retained a single definition for both countries. The Belgian intervals are wider because that subset has fewer events, which could be another reason not to tune the definition separately by country, in our case.

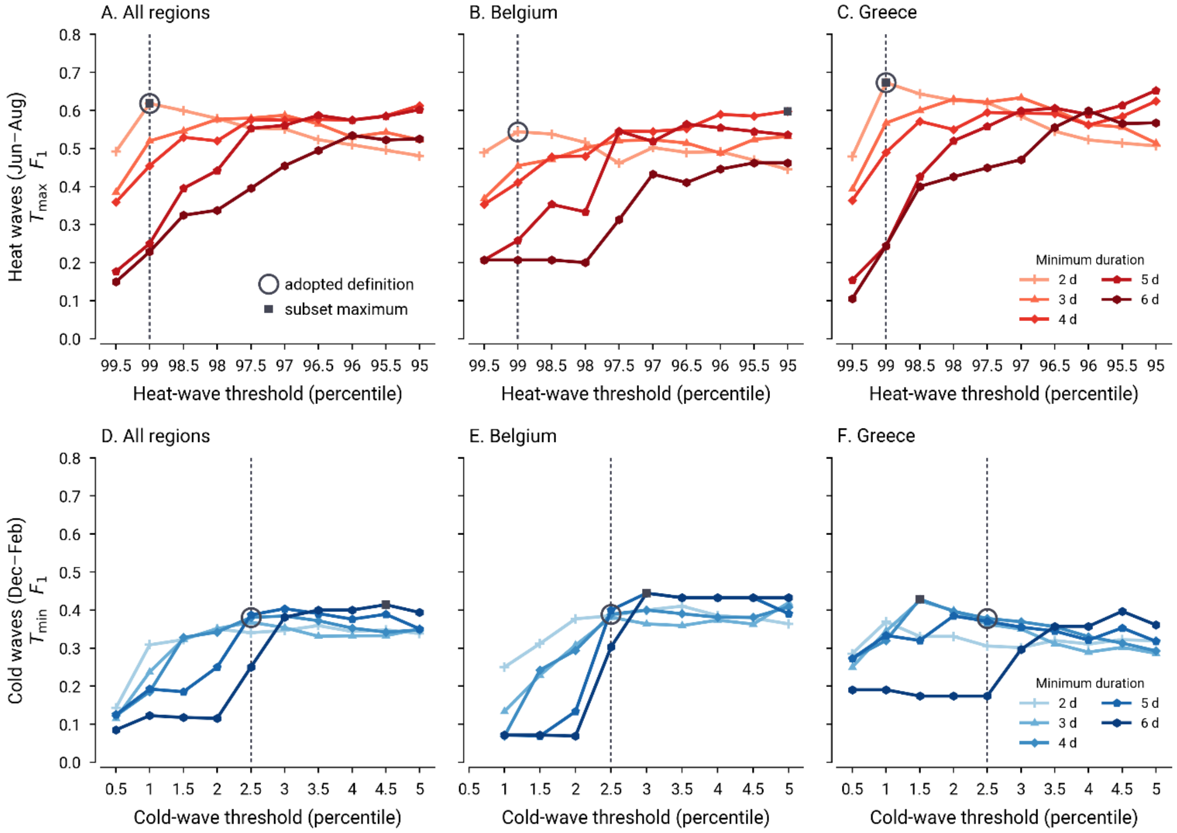


**eFigure 6. Sensitivity of the Coincidence Between Extreme Temperature Events and Excess Mortality Events to the Event Definition, by Country.** Each panel plots the F-measure against the definition threshold, with one line per minimum duration: all six metropolitan areas pooled (A, D), the three Belgian areas (B, E), and the three Greek areas (C, F). Heat waves are shown above and cold waves below, each scored on the variable of its own definition. The dashed vertical line and the open circle mark the adopted definition; the filled square marks the highest-scoring cell of each subset, and coincides with the circle wherever the adopted definition is already optimal. For heat waves the Greek maximum is the adopted definition, while the Belgian nominal maximum reaches 0.60 (95% CI 0.46–0.73) against 0.54 (0.39–0.67) at the adopted definition. Intervals are 95% block-bootstrap intervals resampling metropolitan-area season-years.

**eTable 6. Coincidence Between Extreme Temperature Events (ETE) and Excess Mortality Events (EME) Under the Adopted Definitions.**

| Subset | Hazard | ETEs matched | EMEs matched | P (95% CI) | R (95% CI) | F1 (95% CI) |
|---|---|---|---|---|---|---|
| *Rate baselines* | | | | | | |
| All six areas | HW | 50/78 | 37/62 | 0.64 (0.53-0.75) | 0.60 (0.47-0.72) | 0.62 (0.51-0.71) |
| All six areas | CW | 15/28 | 13/44 | 0.54 (0.32-0.73) | 0.30 (0.17-0.44) | 0.38 (0.23-0.52) |
| Belgium | HW | 19/39 | 16/26 | 0.49 (0.34-0.63) | 0.62 (0.43-0.80) | 0.54 (0.39-0.67) |
| Belgium | CW | 7/11 | 7/25 | 0.64 (0.33-0.91) | 0.28 (0.12-0.44) | 0.39 (0.19-0.56) |
| Greece | HW | 31/39 | 21/36 | 0.79 (0.67-0.90) | 0.58 (0.41-0.74) | 0.67 (0.54-0.78) |
| Greece | CW | 8/17 | 6/19 | 0.47 (0.18-0.73) | 0.32 (0.11-0.57) | 0.38 (0.13-0.60) |
| *Count baselines* | | | | | | |
| All six areas | HW | 49/78 | 36/61 | 0.63 (0.51-0.73) | 0.59 (0.46-0.72) | 0.61 (0.50-0.70) |
| All six areas | CW | 15/28 | 13/39 | 0.54 (0.33-0.73) | 0.33 (0.20-0.49) | 0.41 (0.25-0.56) |

In addition, we ran further checks to confirm the robustness of the reported $F_1$ scores. First, a sensitivity check refitting the baseline on weekly death counts rather than rates leaves them essentially unchanged ($F_1$ 0.61 for heat waves and 0.41 for cold waves). No individual excess mortality event drives them: excluding any one event changes $F_1$ by at most 0.032 for either hazard. And the season rule is consequential for few events — 153 of the 170 excess mortality events (90%) fell wholly within a single season, and assigning events instead by the season holding most of their weeks, by their midpoint week, or admitting them to every season they overlapped changed $F_1$ by at most 0.028 and in no case altered which definitions were preferred. The longest event, in Thessaloniki, spans 41 weeks and all four seasons, so no single-season label describes it; it is retained under its season of onset, and removing it changes $F_1$ by only 0.016.

#### III. Additional Results

##### Excess Mortality per Excess Week

Excess mortality was attributed to temperature extremes on a week-by-week basis rather than on an event-by-event basis. Crediting an entire event's excess to a temperature extreme that overlaps it at any point is harmless for a one-week event and misleading for a long one: the largest excess mortality events in this study run for tens of weeks with a temperature extreme covering only a small fraction of them, and event-level attribution would assign nearly all of their deaths to heat or cold. A week is therefore attributed to a temperature extreme when the extreme overlaps that week or ended within the preceding seven days. This is the linkage rule of Section II read at weekly rather than event resolution: the union of the per-week windows across an event's weeks is exactly the interval used to link events to temperature extremes, so an event is linked to an extreme if and only if at least one of its weeks is attributed to it. The six categories — inside or outside a pandemic wave, crossed with heat wave, cold wave, or neither — are mutually exclusive and partition the 14,210 excess deaths exactly.

eTable 7 reports the result. Outside the pandemic period, an excess-mortality week coinciding with a temperature extreme saw about three times as many excess deaths as one without: 30.8 deaths per week for heat waves and 28.2 for cold waves, compared with 10.5 for weeks with no coincident extreme. During the pandemic period, the contrast disappears, with excess deaths ranging from 41 to 49 regardless of whether a temperature extreme coincided, consistent with pandemic mortality dominating the signal over that interval. Across the study period as a whole, 3,384 of the 14,210 excess deaths (24%) fell in weeks coinciding with a temperature extreme, in 96 of the 456 excess weeks (21%).

"During the pandemic period" denotes temporal overlap with a national COVID-19 wave, not attribution of cause. The Greek wave spans October 2020 to December 2022 and so covers two entire summers, including the 2021 Greek and 2022 European heat waves; excess mortality in that category cannot be apportioned between heat and pandemic causes.

**eTable 7. Excess Mortality per Excess Week, by Pandemic Period and Coincident Temperature Extreme, All Six Metropolitan Areas, 2015–2024.** Weeks are the unit of attribution, so the six categories are mutually exclusive and sum to the total. A week is attributed to a temperature extreme when the extreme overlaps that week or ended within the preceding seven days. "During the pandemic period" denotes temporal overlap with a national COVID-19 wave, not attribution of cause.

| Period | Coincident extreme | Excess deaths | Excess weeks | Deaths per week |
|---|---|---|---|---|
| Outside the pandemic period | Heat wave | 1,294 | 42 | 30.8 |
| Outside the pandemic period | Cold wave | 677 | 24 | 28.2 |
| Outside the pandemic period | No temperature extreme | 1,338 | 128 | 10.5 |
| During the pandemic period | Heat wave | 1,021 | 22 | 46.4 |
| During the pandemic period | Cold wave | 392 | 8 | 49.0 |
| During the pandemic period | No temperature extreme | 9,488 | 232 | 40.9 |
| **Total** | | **14,210** | **456** | **31.2** |

**Annual Heatwave and Cold-Wave Trends**

eFigure 7 shows extreme temperature events over the full 1991–2024 record, aggregating the heatwaves and cold waves detected under the adopted definitions into seasonal series of three metrics per metropolitan area: (i) the number of events, (ii) their cumulative duration in days, and (iii) their total excess temperature, the cumulative °C beyond the threshold on event days. Statistics are reported by the year in which the season starts, so, for instance, the Belgian cold wave of February 2021 is indexed to 2020 as part of the 2020–21 winter. Because the thresholds are fixed percentiles of the 1991–2020 distribution, these series measure exceedance of a fixed absolute threshold rather than changes in relative extremes.

Heat waves became more frequent, longer, and more intense over the record. Comparing the two non-overlapping halves of the record, the six areas together recorded 5.2 heat waves per year over 1991–2014, compared with 7.8 per year over 2015–2024, with cumulative duration rising from 13.1 to 26.0 days per year and mean event duration increasing from 2.5 to 3.3 days. The trend is statistically significant across the whole record (Kendall's $\tau = +0.36$, $p = 0.004$ for the number of events; $\tau = +0.44$, $p < 0.001$ for cumulative duration; $\tau = +0.39$, $p = 0.001$ for excess temperature). It appears in both countries and is proportionally larger in Belgium, where the three areas together move from 2.3 to 3.9 events per year, than in Greece, where they move from 3.0 to 3.9.

Cold waves moved in the opposite direction, but not significantly. Over complete winter seasons, the six areas recorded 4.1 cold waves per year during 1991–2014, compared with 2.9 per year during 2015–2023, with cumulative duration falling from 23.3 to 16.1 days per year. Across the whole record, however, neither trend is distinguishable from zero ($\tau = -0.10$, $p = 0.45$ for the number of events; $\tau = -0.10$, $p = 0.43$ for cumulative duration), so this is better described as a difference between periods than as an established trend. The 1990–91 and 2024–25 winters are excluded because the record runs from January 1991 to December 2024 and neither season is complete.

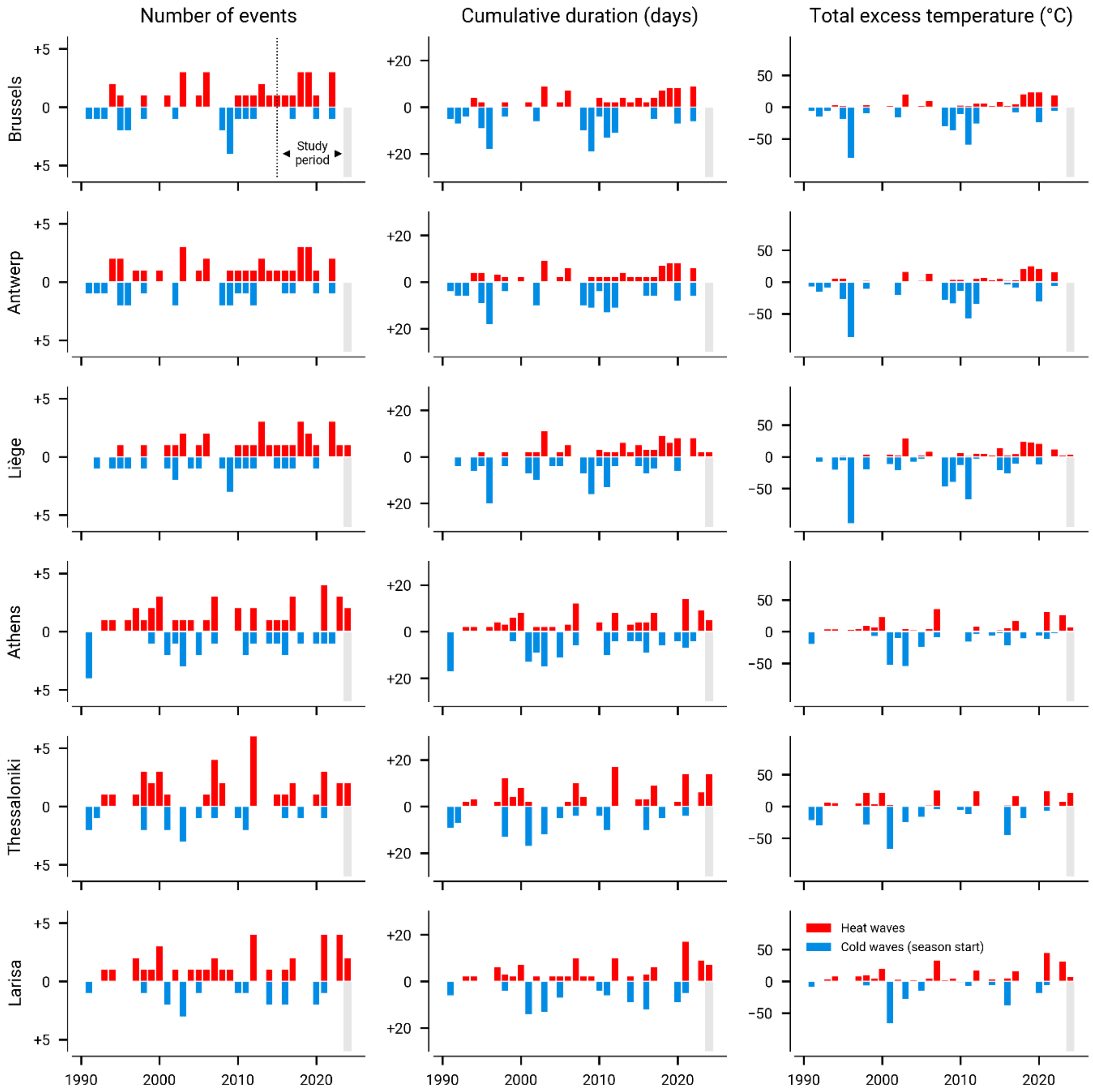


**eFigure 7. Seasonal Trends in Heatwave and Cold-Wave Characteristics per Metropolitan Area, by Season Start Year (1991–2024).** Events are those of the adopted definitions: heat waves of at least two consecutive days with a daily maximum temperature at or above the 99th percentile, cold waves of at least four consecutive days with a daily minimum temperature at or below the 2.5th percentile, both relative to the 1991–2020 climate reference period and computed for each metropolitan area. Columns give the number of events, their cumulative duration in days, and their total excess temperature as the cumulative °C beyond the threshold on event days. Heatwave statistics are in red above the zero line; cold-wave statistics are in blue and mirrored below it, so magnitude increases downward in the first two columns, while cold-wave excess temperature is negative by construction. The x-axis gives the year in which the season starts, so cold-wave statistics cover the winter spanning the displayed year and the next. The shaded band in the lower half of the final year marks the incomplete 2024–25 winter, for which no cold-wave statistics are shown. The dotted vertical line and the annotation in the top-left panel mark the 2015–2024 study period.

**Review of Seasonal Influenza Events**

Seasonal influenza epidemics were reviewed at the national rather than metropolitan-area level, using country-specific sources: end-of-season reports from Sciensano (www.sciensano.be), the Belgian federal institute for health, and from EODY (www.eody.gov.gr), the Greek National Public Health Organization. eTable 8 gives the epidemic period and peak of every season from 2014–2015 to 2023–2024 in both countries.

Influenza surveillance runs from week 40 of one year to week 20 of the following year in both countries, so the surveillance season is identical every year and is not tabulated. Epidemic periods follow the national definition, where the report states one. Belgian reports indicate the weeks when the influenza-like illness epidemic threshold was crossed. Greek reports give the week activity began to rise, and the week it peaked; four of them also give the week the decline ended, and the remaining five end only with a gradual decline running to the close of the surveillance period. For those five marked in the table, the end is the week when activity returns to the highest pre-epidemic level on the report's own surveillance curve. The epidemic peak is the week with the highest influenza activity during the period. Every date is standardized to the Monday of its ISO week, and a period includes both its first and its last week. The 2020–2021 season saw no influenza circulation in either country and is recorded as N.D.

In the main manuscript, Table 3 codes an excess mortality event as coinciding with influenza when the event overlaps the epidemic period of its own country, with both intervals extended to the Sunday that closes their final week.

**eTable 8. Seasonal Influenza Epidemic Periods and Peaks in Belgium and Greece, 2014–2015 to 2023–2024, as Reported by the National Reference Centers.**

| Country | Season | Epidemic start[a] | Epidemic end[a] | Epidemic peak[a] | Ref. |
|---|---|---|---|---|---|
| Belgium | 2014–2015 | 2015-01-12 (W03) | 2015-03-16 (W12) | 2015-02-02 (W06) | [1] |
| Belgium | 2015–2016 | 2016-01-25 (W04) | 2016-03-28 (W13) | 2016-02-29 (W09) | [2] |
| Belgium | 2016–2017 | 2017-01-09 (W02) | 2017-02-20 (W08) | 2017-01-30 (W05) | [3] |
| Belgium | 2017–2018 | 2018-01-08 (W02) | 2018-03-26 (W13) | 2018-03-05 (W10) | [4] |
| Belgium | 2018–2019 | 2019-01-21 (W04) | 2019-03-11 (W11) | 2019-02-11 (W07) | [5] |
| Belgium | 2019–2020 | 2020-01-20 (W04) | 2020-03-09 (W11) | 2020-02-03 (W06) | [6] |
| Belgium | 2020–2021 | N.D. | N.D. | N.D. | [7] |
| Belgium | 2021–2022 | 2022-03-07 (W10) | 2022-04-11 (W15) | 2022-03-21 (W12) | [8] |
| Belgium | 2022–2023 | 2022-12-12 (W50) | 2023-03-20 (W12) | 2022-12-19 (W51) | [9] |
| Belgium | 2023–2024 | 2023-12-11 (W50) | 2024-02-19 (W08) | 2024-01-29 (W05) | [10] |
| Greece | 2014–2015 | 2014-12-15 (W51) | 2015-04-06 (W15)[b] | 2015-01-19 (W04) | [11] |
| Greece | 2015–2016 | 2016-01-04 (W01) | 2016-02-22 (W08) | 2016-02-01 (W05) | [12] |
| Greece | 2016–2017 | 2016-12-26 (W52) | 2017-02-20 (W08) | 2017-01-02 (W01) | [13] |
| Greece | 2017–2018 | 2017-12-25 (W52) | 2018-04-02 (W14)[b] | 2018-02-19 (W08) | [14] |
| Greece | 2018–2019 | 2018-12-24 (W52) | 2019-03-25 (W13)[b] | 2019-02-04 (W06) | [15] |
| Greece | 2019–2020 | 2019-12-30 (W01) | 2020-03-16 (W12)[b] | 2020-02-03 (W06) | [16] |
| Greece | 2020–2021 | N.D. | N.D. | N.D. | [17] |
| Greece | 2021–2022 | 2022-03-07 (W10) | 2022-04-11 (W15)[b] | 2022-03-21 (W12) | [18] |
| Greece | 2022–2023 | 2022-12-12 (W50) | 2023-04-24 (W17) | 2022-12-26 (W52) | [19] |
| Greece | 2023–2024 | 2023-12-11 (W50) | 2024-03-18 (W12) | 2023-12-25 (W52) | [20] |

[a] Dates give the Monday of the corresponding ISO week; a period includes both its first and its last week. Every start and peak week, and every Belgian end week, is stated in the source report. [b] Epidemic end not stated in the report; taken as the week activity returned to the highest pre-epidemic level on the report's own surveillance curve. N.D.: no influenza detected.
